# Transcriptome-wide outlier approach identifies individuals with minor spliceopathies

**DOI:** 10.1101/2025.01.02.24318941

**Authors:** Maggie T. Arriaga, Rodrigo Mendez, Rachel A. Ungar, Devon E. Bonner, Dena R. Matalon, Gabrielle Lemire, Pagé C. Goddard, Evin M. Padhi, Alexander M. Miller, Jonathon V. Nguyen, Jialan Ma, Kevin S. Smith, Stuart A. Scott, Linda Liao, Zena Ng, Shruti Marwaha, Guney Bademci, Stephanie A. Bivona, Mustafa Tekin, Undiagnosed Disease Network, Genomics Research to Elucidate the Genetics of Rare Diseases consortium, Jonathan A. Bernstein, Stephen B. Montgomery, Anne O’Donnell-Luria, Matthew T. Wheeler, Vijay S. Ganesh

## Abstract

RNA-sequencing has improved the diagnostic yield of individuals with rare diseases. Current analyses predominantly focus on identifying outliers in single genes that can be attributed to cis-acting variants within the gene locus. This approach overlooks causal variants with trans-acting effects on splicing transcriptome-wide, such as variants impacting spliceosome function. We present a transcriptomics-first method to diagnose individuals with rare diseases by examining transcriptome-wide patterns of splicing outliers.

Using splicing outlier detection methods (FRASER and FRASER2) we characterized splicing outliers from whole blood for 390 individuals from the Genomics Research to Elucidate the Genetics of Rare Diseases (GREGoR) and Undiagnosed Diseases Network (UDN) consortia. We examined all samples for excess intron retention outliers in minor intron containing genes (MIGs). Minor introns, which make up about 0.5% of all introns in the human genome, are removed by small nuclear RNAs (snRNAs) in the minor spliceosome.

This approach identified five individuals with excess intron retention outliers in MIGs, all of which were found to harbor rare, biallelic variants in minor spliceosome snRNAs. Four individuals had rare, compound heterozygous variants in *RNU4ATAC*, which aided the reclassification of four variants. Additionally, one individual had rare, highly conserved, compound heterozygous variants in *RNU6ATAC* that may disrupt the formation of the catalytic spliceosome, suggesting a novel gene-disease candidate. These results demonstrate that examining RNA-sequencing data for transcriptome-wide signatures can increase the diagnostic yield of individuals with rare diseases, provide variant-to-function interpretation of spliceopathies, and uncover novel disease gene associations.

## MAIN

Rare diseases – those affecting less than 1 in 2,000 people^1,2^ – are collectively common and impact approximately 300 million people globally^3,4^. Despite the high prevalence of rare diseases as a whole, less than 50 percent of individuals with a rare disease have a definitive genetic diagnosis^5–8^. RNA-sequencing (RNA-seq) has proven useful for improving the diagnostic yield by identifying aberrant gene expression or splicing, which refines the search space of potentially causal genetic candidates^9–16^. Such analyses focus primarily on identifying rare variants resulting in single outlier events (i.e., over or under-expression of total gene expression, or aberrant splicing of the canonical isoform). Many studies exclude individuals with many RNA-seq outliers, as technical noise is often thought to be the main driver of this excess^17–22^.

This *cis,* gene-centric approach overlooks single causal monogenic mechanisms that could cause transcriptome-wide outlier splicing patterns, such as pathogenic variants in the spliceosome. Such patterns may be prevalent as the spliceosome consists of over 300 components^23,24^ and two separate snRNA machineries^25,26^: the major spliceosome, which removes ∼99.5% of all introns, and the minor spliceosome, which removes the remaining ∼800 minor introns ^25,27^. Already, many components of the spliceosome have been implicated in human Mendelian diseases^28^, as have many disorders of short-tandem repeat expansions, which sequester vital components of the spliceosome^29,30^. Prior work has shown that pathogenic variants that impact the spliceosome’s function can cause aberrant splicing transcriptome-wide^29–36^. We hypothesized that some individuals with excess splicing outliers have causal variants that impact the splicing process transcriptome-wide, such as variants in the spliceosome.

Some spliceopathies are associated with known transcriptome-wide patterns of aberrant splicing^31–33,37–40^. For example, pathogenic variants in *SF3B1* are associated with intron retention of large introns (>1kb)^33^, while pathogenic variants in *PPIL1* cause retention of short and high GC-content introns^31^ and pathogenic variants in the minor spliceosome snRNA *RNU4ATAC* lead to retention of minor introns^32^.

Given these observations and the increasing availabilityof RNA-seq data in rare disease cohorts, such as the Undiagnosed Diseases Network (UDN) and Genetics Research to Elucidate the Genomics of Rare disease (GREGoR), we investigated an approach to diagnose patients with rare disease by examining patterns in transcriptome-wide splicing outliers. Specifically, we used FRASER^41^ and FRASER2^42^ to examine RNA-seq data from 390 individuals for excess intron retention outliers in minor intron containing genes (MIGs). This approach allowed us to diagnose four individuals with RNU4atac-opathy, provide variant-to-function resolution in these individuals, and uncover a novel, putative gene-disease candidate (*RNU6ATAC*). These results validate the ability of transcriptome-wide outlier splicing patterns to be a useful unbiased diagnostic approach in the analysis of rare disease cohorts.

## METHODS

### Study cohort

#### UDN and GREGoR Stanford sites

We performed RNA-sequencing on 422 whole blood samples from the Genomics Research to Elucidate the Genetics of Rare diseases (GREGoR) and Undiagnosed Diseases Network (UDN) consortia. After removing samples with missing metadata or insufficient RNA quality, we filtered our cohort to 390 whole blood samples. We ran the outlier detection methods FRASER^41^ and FRASER2^42^ on RNA-seq data from our filtered cohort. The primary systems affected for the 390 individuals with rare diseases in our filtered cohort were neurologic (n=91) and musculoskeletal (n=28) **(Figure 1)**. 120 samples were novel and 270 were previously published in Ungar et al., 2024^43^. Ethical and research approvals were provided by the Stanford University IRB (protocol 60837), and the National Human Genome Research Institute Institutional Review Board (IRB) (protocol 15-HG-0130). All participants provided informed consent.

**Figure 1.**
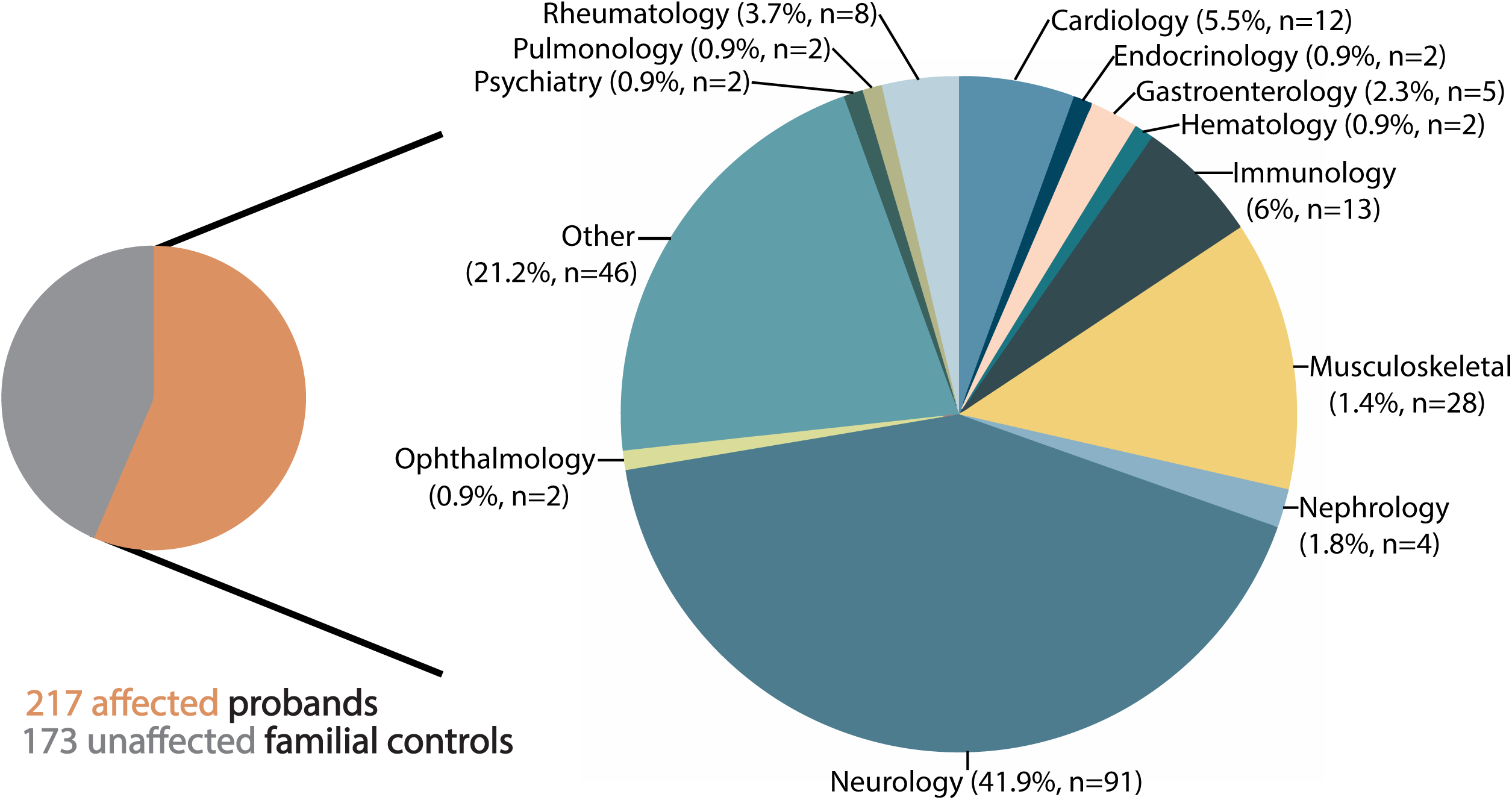
Affected status and primary systems affected for individuals in our GREGoR and UDN cohort. Two pie charts depicting information on our rare disease cohort. The pie chart on the left of the page depicts the affected status composition of our 390-person rare disease cohort from the Undiagnosed Disease Network (UDN) and Genomics to Elucidate the Genetics of Rare Disease (GREGoR) consortia. Lines radiating from the affected (orange) section of the left pie chart indicate that the pie chart on the right contains information on the primary systems affected for all 217 individuals with rare disease in our cohort. The percent and number of individuals affected (n) per system in our cohort are labeled. “Cardiology” includes both cardiology and vascular systems, while “Immunology” includes immunology, infectious diseases, and allergies. “Musculoskeletal” also includes orthopedic complaints. “Other” indicates that no system was selected for the individual at the time of enrollment.

**Figure 2.**
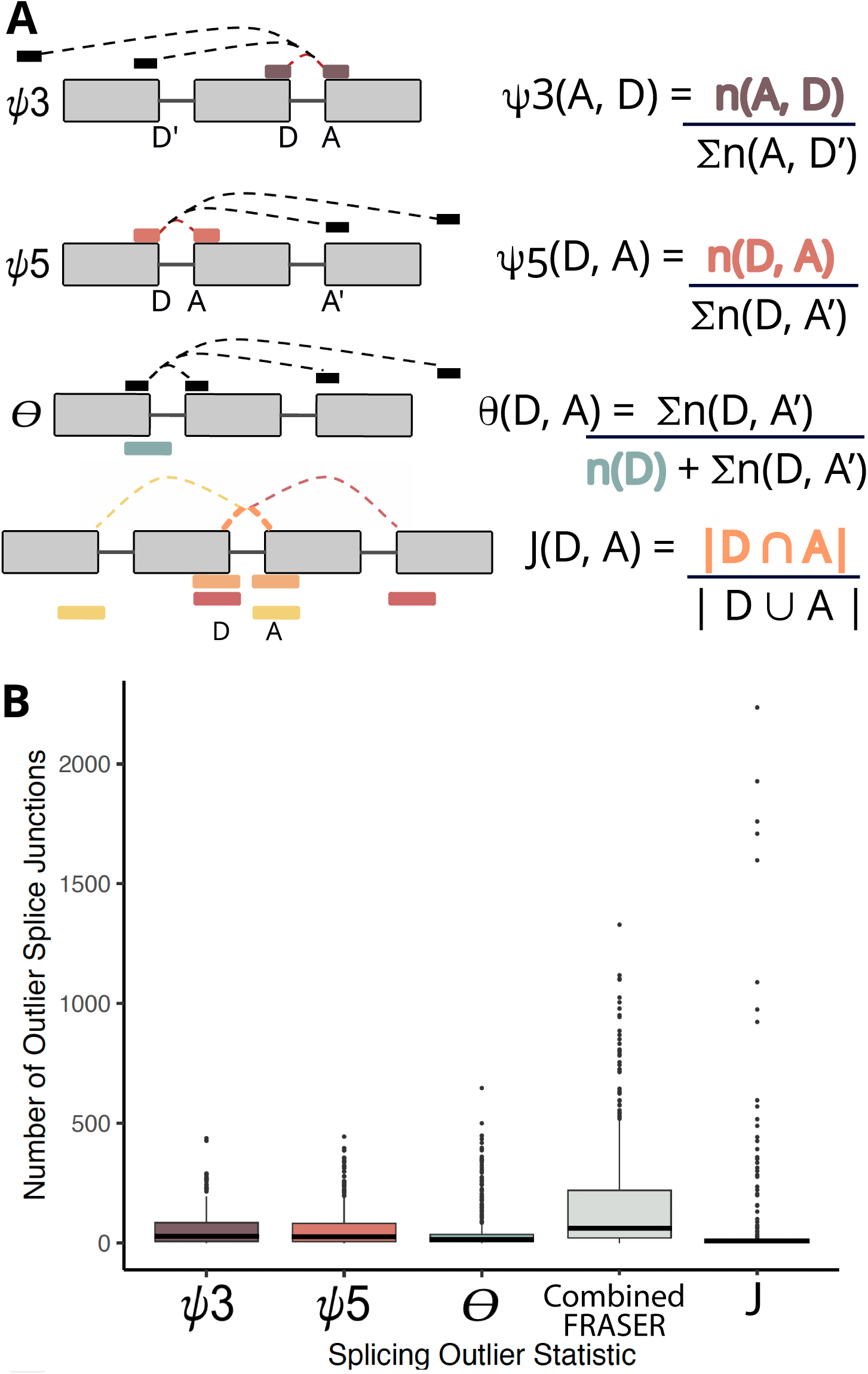
Evaluation of the number of outlier junctions detected per person. (A) Depiction of FRASER and FRASER2 metrics. ψ5 and ψ3 use split reads, which span an exon-exon boundary, while θ and the Jaccard index (J) use both split and unsplit reads, the latter of which span only one exon boundary. ψ5 quantifies alternative acceptor usage from a single donor, while ψ3 quantifies alternative donor usage from a single acceptor. θ quantifies the splicing efficiency, which captures intron retention outliers by comparing the number of split reads to all reads, split and unsplit. J quantifies the proportion of all reads, split and unsplit that support the spicing of an intron of interest compared to all reads, split and unsplit, associated with both the donor and acceptor of the intron of interest. (B) Box plot displaying the number of outlier splice junctions detected per person for each metric. Junctions were labeled as outliers if their adjusted p-value was less than 0.05 and their absolute value of |Δψ|, a normalization metric, was greater than 0.3. The Y-axis is the number of outlier junctions found per person, while the X-axis represents the different types of metrics examined by FRASER (ψ3, ψ5, θ) and FRASER2 (J). Combined FRASER refers to the combined number of outlier junctions from FRASER’s three metrics-ψ5, ψ3, and θ. The bottom of the box represents the first quartile, while the middle line represents the median and the top of the box represents the third quartile. All dots represent outliers.

#### Genome sequencing

The details of clinical and research genome sequencing for cases A1, B1, C1, C2, and D1 are described in **Supplemental Methods**. The *RNU4ATAC* variants in cases A1, C1, and C2 were initially identified by clinical genome sequencing. Clinical genome sequencing for cases B1 and D1 were initially nondiagnostic. Targeted analysis of minor spliceosome related genes in the UDN and GREGoR variant call sets at Stanford led to the identification of the *RNU4ATAC* and *RNU6ATAC* variants in cases B1 and D1, respectively. Confirmation of variants and segregation in parents was performed by Sanger sequencing.

#### Sample collection and preprocessing

RNA-sequencing was performed on whole blood from 422 samples from 422 affected individuals and familial controls. The experimental collection and preprocessing protocol for 287 whole blood samples can be found in Ungar et al., 2024^43^. Of the 287 samples described in Ungar et al., 2024^43^, eight samples were collected and processed in PAXgene tubes at the Utah UDN site before being shipped to Stanford.

For the remaining 135 whole blood samples, 128 were collected and processed in Paxgene RNA tubes at Stanford per the manufacturer’s protocol (PreAnalytix Company). The concentration of isolated total RNA was determined using the QUBIT HS RNA kit (ThermoFisher) and the Bioanalyzer 2100 (Agilent) Nano RNA chip for RIN quality score. Additionally, seven whole blood samples were collected and processed at the Miami UDN site. There, the samples were collected in Paxgene tubes, and the concentration of isolated total RNA and RIN were determined using the Agilent 4200 Tape Station and Qubit 3.0 fluorometer. The RIN was greater than 8 for all 128 samples processed at Stanford per the PreAnalytix Company protocol and greater than 7 for all samples processed at the Miami UDN site.

#### RNA-seq library preparation and sequencing

The experimental protocol for 287 whole blood samples (including the 8 samples from the Utah UDN site) can be found in Ungar et al., 2024^43^. In brief, cDNA libraries were generated using either the Illumina TrueSeq Stranded mRNA Sample Prep Kit protocol and dual indexed or the Universal Plus mRNA-seq NuQuant library prep protocol from Tecan following the same protocol as Amar et al., 2022^44^.

Of the remaining 135 whole blood samples, 128 underwent processing using the following workflow at Stanford, and the remaining 7 were processed at the Miami UDN site. At Stanford, 500ng total input RNA was used for library construction using the Tecan Universal Plus RNA-SEQ with NuQUANT and AnyDeplete Module to remove globin and ribosomal RNAs. Libraries were dual-indexed and also contained a unique molecular index (UMI). The libraries were produced in 96 well plates using a Biomek i7 Liquid Handler robot with Tecan library-specific scripts. Completed cDNA libraries were quantified using the QUBIT HS DNA kit (ThermoFisher) and library tracings were generated using the Advanced Analytical Fragment Analyzer for average base pair length and general integrity. Individual cDNA libraries were normalized to equivalent molar concentration, pooled into batches of 64 samples, and sequenced at low depth 2×100bp using an Illumina iSeq. Pooled libraries were then rebalanced based on reads obtained and each pool was loaded onto an Illumina Novaseq S2 Flowcell, and sequenced as 2×150bp pair-ended reads.

At the Miami UDN site, globin and ribosomal RNAs were removed using the Illumina Ribo-Zero Plus rRNA Depletion Kit. The cDNA libraries were generated using the Illumina Stranded Total RNA Prep, Ligation Kit with Ribo-Zero Plus, dual indexed using IDT for Illumina RNA UD Indexes, and prepped using the Hamilton MicroLab Star liquid handling instrument. As with the samples at the Stanford site, libraries were dual-indexed and contained a UMI. Agilent 4200 Tape Station and Qubit 3.0 fluorometer were then used to determine proper library dilution and balance samples across sequencing runs at the Miami UDN site. Pooled and balanced libraries were then sequenced on an Illumina NextSeq 550, in which 9 and 10 samples were included per flow cell. The Illumina NextSeq 500/550 high Output Reagent V2 Kit (300 cycles, 2 x 150 bp) and Illumina NextSeq 550 High Output Flow Cell V2.5 sequencing kits were used, resulting in all runs generating 150-bp paired-end reads. All reads were filtered according to Illumina recommendations for RNAseq (CPF> 80%, Q>30%>80% for all samples). Specifically, over 80% of the clusters on the flow cell met the Cluster Passing Filter threshold and more than 80% of the total bases sequenced had a Phred quality score of Q30 or higher.

After sequencing, we removed 32 samples from further analysis due to missing information or insufficient RNA quality. Specifically, we removed 14 samples with a RIN less than seven, 14 samples with missing RINs, four samples with missing ages, and two samples with unknown affected statuses from the outlier calling analysis. Our remaining filtered cohort consisted of 217 samples from 217 affected individuals and 173 samples from 173 unaffected familial individuals.

### Pipeline

#### Transcriptome quality control and alignment

The computational pipeline for quality control and alignment of the 270 whole blood samples (including the 8 samples from the Utah UDN site) is described in Ungar et al., 2024^43^. In short, we generated FASTQ files by demultiplexing BCL data using bcl2fastq (https://emea.support.illumina.com/sequencing/sequencing_software/bcl2fastq-conversion-software.html), then reads were aligned to the hg38 human reference genome using STAR (2.8.4a)^45^ and the GENCODEv35^46^ primary genome annotations. Adapters were removed and reads were trimmed using cutadapt (version=2.4)^47^ (https://github.com/marcelm/cutadapt) and reads with a mapping quality under 30 were removed.

FASTQ files for the additional 120 whole blood samples, processed at both Stanford (n=113) and the Miami UDN site (n=7), were also generated by demultiplexing using bcl2fastq. We aligned the reads with the hg38 human reference genome with STAR (version=2.7.10a)^45^, as well as the GENCODEv39^46^ primary genome annotations. As all 120 files had a read length of 150, an annotation file for STAR/2.7.10a was created for an overhang of 100. The adapters were removed and reads trimmed using cutadapt (version=2.4)^47^ (https://github.com/marcelm/cutadapt) with a minimum trim length of 50. All reads with a mapping quality less than 30 were removed, optical duplicates were filtered from the aligned bams using Picard (http://broadinstitute.github.io/picard), and a minimum pixel distance of 2500 px was set.

#### Splicing outlier calling

To assess splicing outliers, we processed all 390 aligned bam files through the first iteration of FRASER (version=1.14.0), which we will refer to as FRASER^41^, and the second iteration of FRASER (version=1.99), which we will refer to as FRASER2^42^. At the filterExpressionAndVariability step, the minimum read count in at least one sample was set at 20 and the minimum ΔѰ reported was set at 0.0. Junctions were defined as significant if their adjusted p-value (q) after false discovery rate (FDR) correction was less than 0.05 and their |ΔѰ| was greater than or equal to 0.3. The pipeline and conda environments for this analysis can be found at https://github.com/maurermaggie/Transcriptome_Wide_Splicing_Analysis/tree/main/FRASER_snakemake.

#### Excess outlier detection

Samples were determined to have an excess of outliers across six metrics: all FRASER^41^ outliers, Ѱ3 outliers, Ѱ5 outliers, *θ* outliers, FRASER2^42^’s Jaccard index outliers, and *θ* outliers within minor intron containing genes (MIGs). In each case, the excess outliers were detected by calculating the mean and standard deviation of the number of significant outlier junctions of their respective type. Samples with a number of outlier junctions greater than two standard deviations from the mean were deemed to have an “excess outlier” status.

We used logistic regression to examine if excess outliers using one metric of FRASER^41^ (Ѱ3, Ѱ5, *θ*, and all combined FRASER^41^ outliers) or FRASER2^42^ (Jaccard index) were significantly more likely to be excess outliers using a different metric. In order to perform this analysis, samples were given the value of 1 if they were an excess outlier in a given metric, and 0 if they were not. For each metric, we used the glm function (family = ‘binomal’, link = ‘logit’) found in R (v4.3.1)^48^ to perform a logistic regression for excess outlier status of all other metrics; for example, Ѱ3 excess outlier status ∼ Ѱ5 excess outlier status + *θ* excess outlier status + FRASER^41^ combined excess outlier status + Jaccard index excess outlier status. FDR correction was applied to all resulting p-values across all tests.

#### Gene set enrichment

Gene set enrichment was run by comparing the enrichment of the non-outlier genes and outlier genes detected by FRASER^41^ and FRASER2^42^. Notably, the enrichment analyses were run separately on the data provided by FRASER^41^ and FRASER2^42^. In both analyses, non-outlier genes were detected by examining the results of the filterExpressionAndVariability step in either FRASER^41^ or FRASER2^42^. This step removes junctions with less than 20 reads in one sample and requires at least one sample have a |Δψ| of 0. We then subsetted the genes detected at this step into those found as outliers in any of our 390 samples, deemed outliers, and those not, which we labeled as non-outliers (or inliers).

We evaluated the enrichment or depletion of outlier genes in multiple gene sets analyzed in Cormier et al., 2022^49^. We examined haploinsufficiency by looking for enrichment of our outlier genes in a set of 294 genes determined to be dosage-sensitive in the ClinGen dataset^50^. In addition, we examined sets of 1183 and 709 genes shown to follow, respectively, an autosomal recessive inheritance pattern or autosomal dominant inheritance pattern by Blekhman et al., 2008^51^ and Berg et al., 2013^52^. As a negative control, we used a set of 371 Olfactory Receptor genes from Mainland, et al., 2015^53^. Since the majority of olfactory genes are known to contain single exons and, thus, not predicted to undergo splicing^54^, we also used CRISPR non-essential genes from Hart et al., 2017^55^ as a second negative control. The final gene set we used from Cormier et al., 2022^49^ is a list of 2072 genes collected across several Deciphering Developmental Disorders studies^56–61^, which are hosted on Gene2Phenotype^56^. We also evaluated the enrichment of outlier genes compared to the complete set of OMIM genes^62^. The location and sources for all of the aforementioned gene sets can be found in **Table S1**.

To examine the correlation between outlier status and enrichment in the above gene sets, we created contingency tables consisting of the following (clockwise from top left): (1) number of genes that are splicing outliers and in the gene set of interest, (2) number of genes that are splicing outliers and not in the gene set of interest, (3) number of genes that are splicing inliers and not in the gene set of interest, and (4) number of genes that are splicing inliers and in the geneset of interest. We then used the fisher.test function found in R (v4.3.1)^48^ to run Fisher’s exact test on contingency tables for each gene set, followed by FDR correction across all of the gene set enrichment analyses.

#### Correlation with technical and biological covariates

We used the lm function found in R (v4.3.1)^48^ to run a linear regression on the correlation between the number of significant outlier splice junctions per person and four technical and biological covariates: RIN, age, sex, and batch. We applied this analysis to both the total number of outlier junctions found using FRASER^41^ and FRASER2^42^.

In addition, we used the glm function (family = ‘binomal’, link = ‘logit’) found in R (v4.3.1)^48^ to run a logistic regression to examine the association between excess outlier status and the same four covariates: RIN, age, sex, and batch. To define outlier status, we assigned samples the value of 1 if they were excess outliers, defined as a sample with a number of significant outlier junctions more than two standard deviations from the mean, and the value of 0 if they were not an excess outlier. For our analysis of the covariates and FRASER^41^ excess outlier status, an individual was marked as an excess outlier if they were an excess outlier across any metric of FRASER^41^ (Ѱ3, Ѱ5, *θ*), as well as if they had an excess total number of significant outliers detected by FRASER^41^ as a whole (combining the total outliers of all three metrics). Since FRASER2^42^ outputs only the Jaccard index metric, samples were marked with a 1 if they contained an excess number of significant Jaccard index outliers.

#### Analysis of intron retention outliers in minor intron containing genes (MIGs)

We sought to examine if an RNA-seq outlier based technique could identify and diagnose individuals with rare, pathogenic, biallelic variants in the minor spliceosome snRNAs *RNU4ATAC* and *RNU12.* To accomplish this, we conducted two analyses examining (1) the number of significant intron retention outliers in MIGs and (2) the number of MIGs impacted by significant intron retention outliers.

To perform these analyses, we filtered all junctions detected by FRASER^41^ to only those that were significant (q < 0.05 and |Δψ| >= 0.3) and of the type *θ* which represents splicing efficiency outliers, which captures partial or full intron retention. Using the list of all significant intron retention outliers per person, we next filtered this list to only MIGs, which we defined as genes found in the Minor Intron Database from Olthof et al., 2019^26^. Specifically we used the Homo_sapiens_intron.csv, which can be downloaded at https://midb.pnb.uconn.edu/return_downloads.php?species_var=Homo+sapiens. We then calculated the number of significant intron retention outliers found in MIGs per individual and the number of MIGs with significant intron retention per individual.

#### Gene set comparison of MIGs with intron retention

To examine the correlation between the MIGs with significant intron retention outliers found between all four RNU4atac-opathy cases (A1-C2), we created contingency tables for each possible case pair: A1-B1, A1-C1, A1-C2, B1-C1, B1-C2, C1-C2. To create the contingency tables, we first created a list of all MIGs with significant intron retention outliers found in the 390-person cohort (here, called the ‘all’ list). We also had two lists of the MIGs with significant intron retention outliers in Sample 1, called ‘sample1’, and MIGs with significant intron retention outliers in Sample 2, called ‘sample2.’ We then created two ‘negative’ lists consisting of the genes in the list of ‘all’ list not present in the ‘sample1’ and the genes in the ‘all’ list not present in the ‘sample2’, which we called ‘not_sample1’ and ‘not_sample2’, respectively.

We used these lists to create a contingency table, which consisted of (going clockwise from the top left): (1) the number of genes shared between the ‘sample1’ and ‘sample2’, (2) the number of genes in ‘sample1’ and ‘not_sample2’, (3) the number of genes in ‘not_sample1’ and ‘not_sample2’, and (4), the number of genes in ‘not_sample1’ and ‘sample2’. To examine the correlation between the MIGs with significant intron retention outliers between two samples, we used the fisher.test function found in R (v4.3.1)^48^ to run a Fisher’s exact test, followed by FDR correction across all tests.

To examine if the MIGs with significant intron retention outliers found in the sample with rare, biallelic variants in *RNU6ATAC* (D1) are shared with the four RNU4atac-opathy cases, we combined the MIGs with significant intron retention outliers from A1-C2 into one list (‘RNU4atac-opathy’), leaving two lists: ‘D1’ and ‘RNU4atac-opathy’. We next created a list of all MIGs with intron retention outliers found in the 390-person cohort (the ‘all’ list) and created two negative lists: first, called ‘not_D1’, which consisted of all of the genes in the ‘all’ list not present in ‘D1’, and second, called ‘not_RNU4atac-opathy’, which comprised of all of the genes in ‘all’ not present in all four of the RNU4atac-opathy cases.

These lists were then used to create a contingency table consisting of (going clockwise from the top left): (1) the number of genes shared between ‘RNU4atac-opathy’ and ‘D1’, (2) the number of genes in ‘RNU4atac-opathy’ and ‘not_D1’, (3) the number of genes in ‘not_RNU4atac-opathy’ and ‘not_D1’, and (4) the number of genes in ‘not_RNU4atac-opathy’ and ‘D1’. We then again used the fisher.test function found in R (v4.3.1)^48^ to run a Fisher’s exact test.

#### Sample size comparison

To examine the portability of our results to researchers and clinicians with access to fewer samples, we created six datasets consisting of 25, 50, 100, 150, 200, 300, and 400 samples. Each dataset was iterative, meaning the samples in each subset are found in all subsets larger than themselves; for example, the samples in the dataset of 50 samples are found in the datasets containing 100, 150, 200, 300, and 400 samples. Each dataset contained no more than one sample with rare, biallelic variants in minor spliceosome snRNAs (A1 for each dataset). Of note, 25 is the smallest number of samples for which FRASER^41^ can generate an output model^41,42^.

We used the lm function found in R (v4.3.1)^48^ to run linear regressions on the sample size and, separately, the number of significant intron retention outliers in MIGs and the z-score of the number of significant intron retention outliers in MIGs.

#### Variant classification

Variants were classified according to the ACMG/AMP criteria^63^ and following the recommendations from Ellingford et al., 2022^64^. Specifically, we applied PM2_Supporting (PM2_P) to variants that are absent or rare (frequency <1% and no homozygous occurrences) in gnomAD v4.1.0; PM3 to variants detected in trans with a pathogenic variant; PS3 when RNA-seq data analyses supported pathogenicity; and PM1 to variants located in mutational hotspots within *RNU4ATAC*, specifically at chromosome 2 (GRCh38)g.121,530,882–121,530,898 and g.121,530,996–121,531,003. Based on these criteria, variants located within the critical regions were classified as pathogenic, while those occurring outside of the critical regions were classified as likely pathogenic.

The interpretation of novel candidate gene-disease relationships was guided by the ClinGen framework^65^. Additional evidence, including more cases and functional studies, is necessary to establish *RNU6ATAC* as a disease-associated gene, which is required before any variant in *RNU6ATAC* can be classified as pathogenic. Therefore, variants found in *RNU6ATAC*, which is considered a “gene of uncertain significance”, were considered candidates and thus classified as “variants of uncertain significance” and reported as “variants in a gene of uncertain significance”.

## RESULTS

### Splicing outliers detected in blood

We hypothesized that global patterns of transcriptomic outliers could inform molecular diagnoses of individuals with rare diseases without *a priori* knowledge of causal variants. In order to address this hypothesis, we analyzed RNA-sequencing on whole blood from a rare disease cohort of 217 affected cases and 173 familial control samples **(Figure 1).** 311 individuals were from the UDN, 48 individuals were from the GREGoR consortium, and 31 individuals were enrolled in both consortia **(Methods)**.

To identify splicing outliers, we used two versions of FRASER: FRASER (version=1.14.0) from Mertes et al, 2021, which we refer to as FRASER^41^, and FRASER (version=1.99) from Scheller et al., 2023, which we refer to as FRASER2^42^. The output of FRASER2^42^ provides a single metric for splicing outliers (Jaccard index)^42^, whereas FRASER^41^ provides more granular metrics for types of splicing outliers (*θ*, Ѱ3, and Ѱ5)^41^, and both outputs informed our analysis. Ѱ3 and Ѱ5 only consider split reads, which are reads that span an exon-exon boundary. Ѱ3 quantifies the alternative donor usage from a single acceptor, or the proportion of reads from an acceptor to a donor of interest compared to the number of reads from the acceptor to all possible donors. Ѱ5 similarly measures the acceptor usage from a single donor, or the number of reads from a donor to an acceptor of interest compared to the number of reads from the donor to all possible acceptors^41^. *θ* and the Jaccard index also utilize unsplit reads, which are reads that do not span an exon-exon boundary of interest^41,42^. The *θ* metric quantifies splicing efficiency outliers, or intron retention outliers, and is calculated by dividing the number of split reads by the number of all reads, split and unsplit, across a donor or acceptor of interest^41^. Finally, FRASER2^42^’s Jaccard index quantifies the proportion of split and unsplit reads supporting the splicing of an intron of interest compared to all reads, split or unsplit, associated with either splice site on the intron of interest^42^ **(Figure 1A)**.

FRASER^41^ quantified 185,484 junctions from 13,137 genes, while FRASER2^42^ identified 236,416 junctions from 17,918 genes across these 390 samples. We filtered the splice junctions to only those with an adjusted p-value (q) less than 0.05 and a |Δψ|, which is a normalization metric, greater than 0.3. We refer to reads passing these filters as significant outlier junctions. On average, we identified a median of 24.5 (IQR [7 - 84.8]) significant outlier Ѱ3 junctions, 25.0 (IQR [6-81.8]) significant outlier Ѱ5 junctions, 11 (IQR [5.3 - 36]) significant *θ* junctions, and 5 (IQR [2-10]) significant outlier Jaccard index junctions per person **(Figure 1B)**.

### Splicing outlier analysis identifies aberrant junctions relevant to rare disease

After detecting outliers, we followed the protocol from Cormier et al., 2022^49^ to identify the relationship between our splicing outlier genes and genes with high or low tolerance to putative loss of function variants. We examined the enrichment of our outlier genes in a set of 294 genes determined to be haploinsufficient in the ClinGen dataset^50^. Olfactory genes are known to be tolerant to putative loss of function variants^54^ so we included known olfactory receptor genes from Mainland et al., 2015^53^ as a negative control. Since the majority of olfactory genes are known to contain single exons and, thus, not predicted to undergo splicing^54^, we also used CRISPR non-essential genes list from Hart et al., 2017^55^ as a second negative control. To examine splicing outliers for their enrichment or depletion within genes with curated disease-causing inheritance patterns, we used genes curated as autosomal dominant and autosomal recessive by Blekman et al., 2008^51^ and Berg et al., 2013^52^. Finally, we investigated the enrichment of our outlier genes in blood in multiple rare-disease relevant gene sets including the complete set of OMIM genes^62^ and genes found to be related to developmental delay and intellectual disability by several Deciphering Developmental Disorders studies^56–61^.

After adjusting the p-value for the FDR, we observed that genes detected as outliers by FRASER^41^ in blood were significantly enriched for OMIM genes (Fisher’s exact test, q=1.47e-40, odds ratio=1.81) and genes implicated in developmental delay and intellectual disability (Fisher’s exact test, q=8.0e-13, odds ratio=1.57). We also found a significant depletion of our splicing outlier gene set for CRISPR non-essential genes (q=0, odds ratio=0) and olfactory receptor genes (Fisher’s exact test, q=0.004, odds ratio=0.11). While significant before correction, there was no significant enrichment for haploinsufficient genes after FDR correction (Fisher’s exact test, q=0.06, odds ratio=1.48). There was also no significant association between splicing outliers and autosomal dominant or recessive genes **(Figure 3A)**.

**Figure 3.**
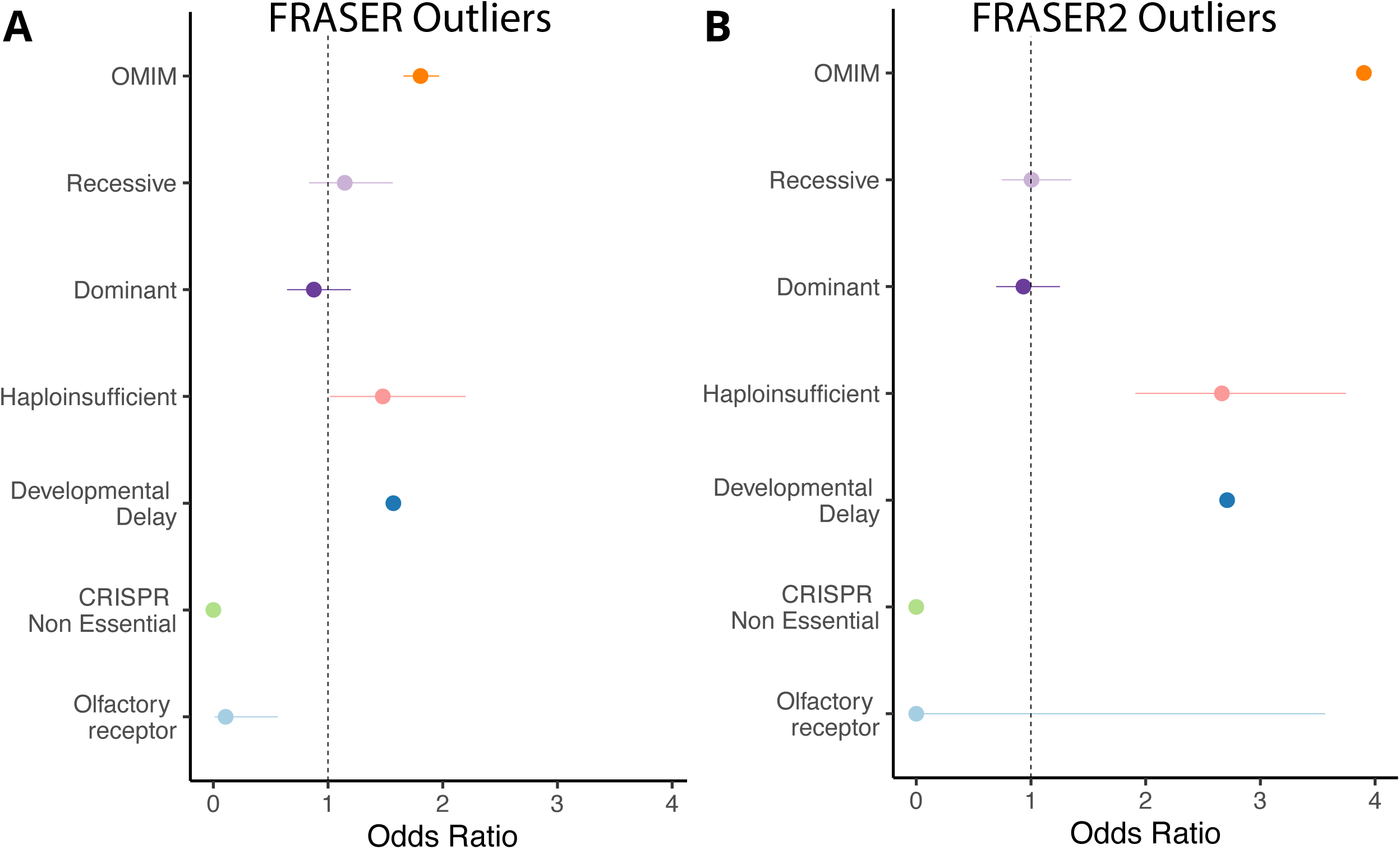
Enrichment analysis on genes detected to be splicing outliers via FRASER1 and FRASER2. (A) Results of enrichment analysis on genes detected to be significant splicing outliers via FRASER. All categories not crossing the dotted vertical line were found to be significant before False Discovery Rate (FDR) correction. All categories shown as significant stayed significant after FDR correction except for the haploinsufficient gene set, whose p-value after correction was 0.06. (B) Results of enrichment analysis on genes detected to have significant Jaccard index outlier junctions (as detected by FRASER2). All categories whose lines do not cross the dotted vertical line were found to be significant before FDR correction, and all categories shown as significant in this figure remained significant after FDR correction.

The enrichment analyses of genes detected as outliers by FRASER2^42^ yielded similar results. There was a significant enrichment of FRASER2^42^ outlier genes for OMIM disease genes (Fisher’s exact test, q=0, odds ratio=3.9) and genes implicated in developmental delay and intellectual disability (Fisher’s exact test, q=7.38e-75, odds ratio=2.71). There was also a similar depletion of CRISPR non-essential genes (q=0, odds ratio=0) and no significant association between FRASER2^42^ outlier genes and autosomal dominant or recessive genes. Unlike the analysis on FRASER^41^ outlier genes, the FRASER2^42^ outlier genes were enriched for haploinsufficient genes after FDR correction (Fisher’s exact test, q=2.29e-9, odds ratio=2.66) and there was no significant association between FRASER2^42^ outlier genes and olfactory receptor genes **(Figure 3B)**.

### Identifying individuals with excess splicing outliers

Our analyses sought to evaluate if excess splicing outlier patterns could be used to increase the diagnostic yield of individuals with rare diseases. To examine the proportion of samples with excess outlier splicing across the transcriptome, we counted the number of significant (q < 0.05 and |Δψ| >= 0.3) outlier splice junctions per individual, **Methods**). We identified individuals as having an excess number of splicing outliers, thus given the status of “excess outliers”, if they had a number of significant outlier junctions greater than or equal to two standard deviations from the mean. Using this method, we found individuals with excess outlier status across all three metrics of FRASER^41^ (Ѱ3, Ѱ5, and *θ*) and FRASER2^42^’s Jaccard index. We found 24 excess Ѱ3 outliers, 24 excess Ѱ5 outliers, 26 excess *θ* outliers, 24 excess FRASER^41^ junction outliers, and 10 excess Jaccard index outliers, **(Figure 4A-D)**. Overall, 40, or 10.3%, of our cohort were found to be excess outliers across at least one metric of FRASER^41^ or FRASER2^42^. Individuals who were excess outliers using one metric, such as Ѱ3, were not more likely to be excess outliers using a different metric, such as Ѱ5, *θ*, or the Jaccard index **(Figure S1)**.

**Figure 4.**
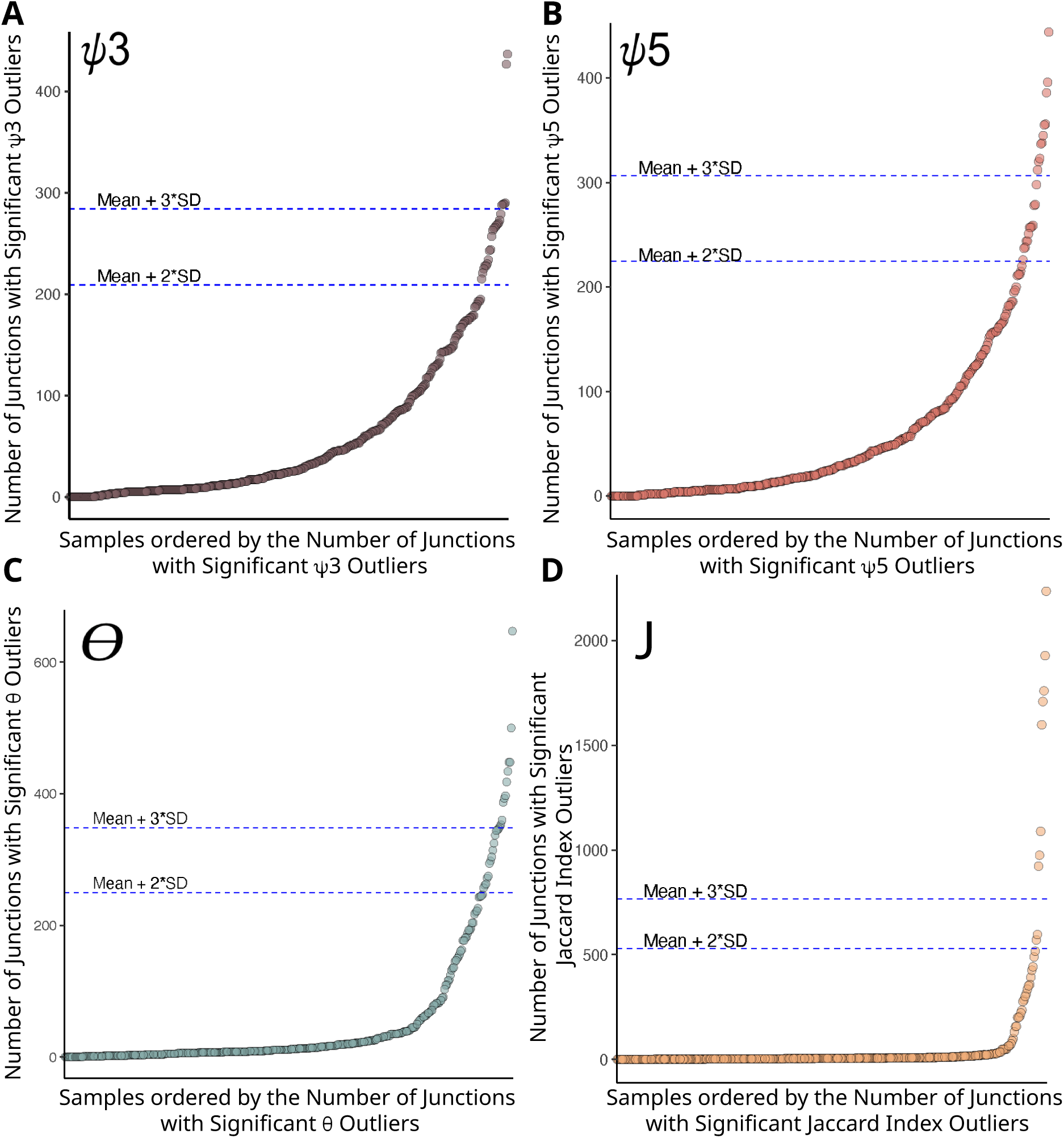
Individuals with excess significant outlier junctions detected using FRASER and FRASER2 metrics. Plots showing the number of significant outlier junctions detected per individual for all three FRASER metrics (ψ3, ψ5, and θ) and the FRASER2 Jaccard index (J). Junctions were labeled as significant outliers if their adjusted p-value was less than 0.05 and their |Δψ|, a normalization metric, was greater than 0.3. For all four plots, each dot represents an individual and the position on the Y-axis of the dot represents the number of significant outlier junctions of the specified type for that individual. Individuals are ordered on the X-axis by the number of significant outlier junctions of the specified metric. The number of significant outliers shown per individual are, as labeled: (A) ψ3, (B) ψ5, (C) θ, and (D) J.

### Correlation between excess outliers and covariates

Rare disease research examining transcriptome outliers has typically excluded samples with excess outliers under the assumption they are driven by technical artifacts^17–22^. We hypothesized that the excess of outliers in at least some samples is driven by biological processes, such as variants impacting the function of the spliceosome.

To evaluate the impact of technical and biological covariates on the number of all total outlier splice junctions per person as calculated by FRASER^41^ (Ѱ3, Ѱ5, and *θ* combined) and FRASER2^42^, we examined the correlation between the number of all outlier splice junctions and RNA integrity number (RIN), sex, batch, and age. The variance in the number of all outlier splice junctions per individual detected by FRASER^41^ was contributed to by RIN (6.0%), sex (63.0%), batch (18.5%), and age (3.5%). Batch was the only term with a significant relationship to the number of significant outlier junctions detected by FRASER^41^ per person (linear regression and FDR correction, q = 1.8e-08, **Figure S2A**). Using FRASER2^42^, we found that the variance in the number of all outlier splice junctions detected per individual was contributed to by RIN (8.7%), sex (3.3%), batch (0.1%), and age (0.1%). There were no significant relationships between the covariates and the number of all outlier splice junctions detected by FRASER2^42^ **(Figure S2A)**.

We next examined the impact of the aforementioned technical and biological covariates on whether an individual was an excess outlier for outlier splice junctions, as determined by FRASER^41^ and FRASER2^42^. Here, individuals were deemed to be a FRASER splice junction excess outlier if they were an Ѱ5 excess outlier, Ѱ3 excess outlier, or *θ* excess outlier, as well as if they were an excess outlier when outliers of all three FRASER^41^ metrics (Ѱ3, Ѱ5, and *θ*) were summed together. For the FRASER^41^ statistics, the covariates contributed to a small portion of the observed variance in excess outlier status. RIN contributed 30.5%; sex, 4.3%; batch, 4.6%; and age, 0.8%. There was a significant relationship between batch and FRASER^41^ outlier status (logistic regression and FDR correction, q=0.003, **Figure S2B**). We also analyzed the relationship between Jaccard index outlier status and the four aforementioned covariates. We found that RIN contributes to the variance in Jaccard index excess outlier status 21.6%; sex, 0.9%; batch, 1.6%; and age 0.7%. There were no significant relationships between the covariates and excess outlier status as detected by FRASER2^42^ **(Figure S2B)**. These results indicate that technical and biological covariates, such as batch, can have significant influences on the quantification of excess outliers and need to be considered when quantifying excess outlier events.

### Cases with excess significant intron retention outliers in MIGs identified as having RNU4atac-opathy

Minor introns make up about 0.5% of all introns in the human genome^66,67^. They diverge from major introns by their splicing motifs and the snRNAs in their spliceosomal machinery^68,69^. Pathogenic variants in two components of the minor spliceosome, *RNU4ATAC* and *RNU12,* cause autosomal recessive Mendelian diseases and have been associated with increased minor intron retention events and aberrant splicing events around minor introns^32,37^. Based on these observations, we examined our cohort’s RNA-seq data for significant *θ* outliers (i.e.intron retention outliers) occurring in minor intron containing genes (MIGs) as defined by the Minor Intron Database from Olthof et al., 2019^26^. Specifically, we examined the number of significant (q<0.05, |Δψ| = 0.3) intron retention outliers in MIGs, as well as the number of MIGs with significant intron retention outliers, per person. The mean number of significant intron retention outliers in MIGs was 5.8, while the median was 0 (IQR [0-2]) per individual **(Figure 5A)**.

**Figure 5.**
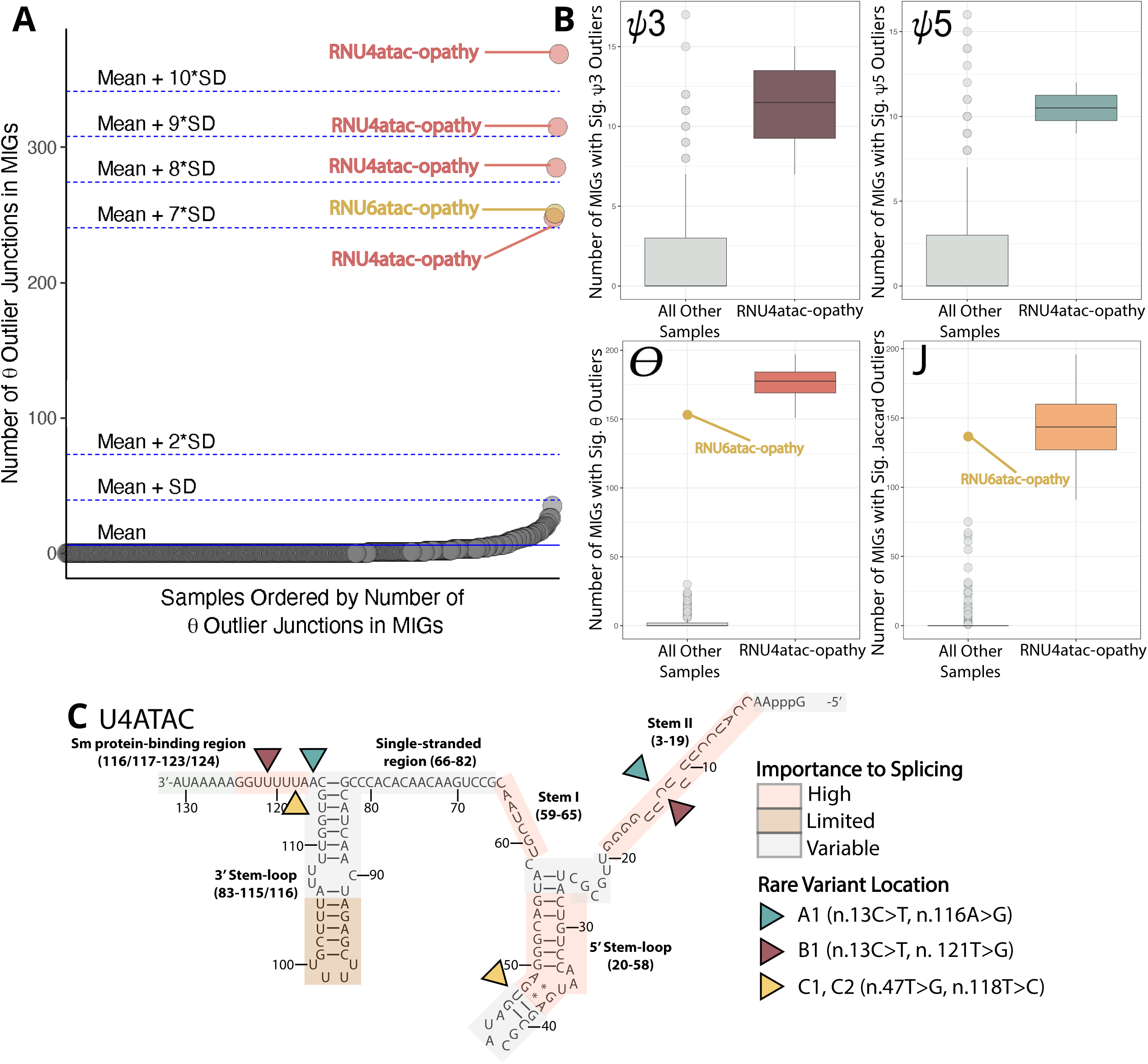
Outliers with excess intron retention events (θ) in minor intron containing genes (MIGs) identified in individuals with rare, biallelic variants in *RNU4ATAC*. (A) Plot showing the number of significant intron retention (θ) events in minor intron containing genes (MIGs). Each dot represents an individual and the Y-axis position represents the number of significant intron retention (θ) events in MIGs detected in that individual. The X-axis is ordered by the number of significant intron retention events (θ) in MIGs. Individuals with rare, biallelic variants in *RNU4ATAC* or *RNU6ATAC* are labeled as “RNU4atac-opathy” and “RNU6atac-opathy”, respectively. (B) Boxplots showing the number of MIGs with significant outliers of type (from top left clockwise) ψ3, ψ5, Jaccard index (J), and θ (intron retention). In each boxplot, the right box labeled “RNU4atac-opathies” represents the individuals with rare, biallelic variants in *RNU4ATAC*, while the left box represents the remaining samples. Where D1 is an outlier, it is marked as an orange circle. Note that the y-axis ends at around 15 for the ψ3 and ψ5 boxplots and around 200 for the θ and J boxplots. (C) Secondary structure of U4ATAC, which is encoded by *RNU4ATAC*. Areas of high importance to splicing are labeled in pink; limited importance to splicing, brown; and variable importance to splicing, gray. The rare variants in individuals A1, B1, C1, and C2 are labeled by the blue, red, and yellow arrows.

Using this method, we identified 5 individuals with an excess of significant intron retention outliers in MIGs (samples A1, B1, C1-2, and D1, **Tables 1-2**). These individuals had a considerable excess of MIGs with significant intron retention outliers (z-scores of 9.2, 8.3, 10.8, 7.2, and 7.3 respectively, **Figure 5A**). This corresponds to 180, 175, 197, 197, and 153 out of 770 possible MIGs impacted, respectively^26^. Notably, A1-D1 did not have an excess of MIGs impacted by outliers in FRASER^41^’s other metrics – Ѱ3 or Ѱ5 – and the pattern of excess aberrant splicing in these samples is limited to significant intron retention outliers in MIGs **(Figure 5B).**

**Table 1.** Genetic information for samples A1-D1. Information on the genetic variants found in *RNU4ATAC* in samples A1-C2, as well as the variants found in *RNU6ATAC* in sample D1. The gene names are italicized according to HGNC standards.

| Family-Case ID | Gene | Genomic coordinates (GRCh38) | HGVS | ACMG Classification | ACMG Criteria | Zygoty | Inheritance Pattern |
| --- | --- | --- | --- | --- | --- | --- | --- |
| **A1** | *RNU4ATAC*<br>(NR_023343.1) | 2-121530892<br>-C-T | n.13C>T | Pathogenic | PS3; PM1;<br>PM2_P; PM3 | Compound<br>Heterozygous | Autosomal<br>recessive |
|  |  | 2-121530995<br>-A-G | n.116A>G | Pathogenic | PS3; PM1;<br>PM2_P; PM3 |  |  |
| **B1** | *RNU4ATAC*<br>(NR_023343.1) | 2:121530892-C-T | n.13C>T | Pathogenic | PS3; PM1;<br>PM2_P; PM3 | Compound<br>Heterozygous | Autosomal<br>recessive |
|  |  | 2-121531000<br>-T-G | n.121T>G | Pathogenic | PS3; PM1;<br>PM2_P; PM3 |  |  |
| **C1** | *RNU4ATAC*<br>(NR_023343.1) | 2-121530926<br>-T-G | n.47T>G | Likely<br>Pathogenic | PS3, PM2_P;<br>PM3 | Compound<br>Heterozygous | Autosomal<br>recessive |
|  |  | 2-121530997<br>-T-C | n.118T>C | Pathogenic | PS3; PM1;<br>PM2_P |  |  |
| **C2** | *RNU4ATAC*<br>(NR_023343.1) | 2-121530926<br>-T-G | n.47T>G | Likely<br>Pathogenic | PS3, PM2_P;<br>PM3 | Compound<br>Heterozygous | Autosomal<br>recessive |
|  |  | 2-121530997<br>-T-C | n.118T>C | Pathogenic | PS3; PM1;<br>PM2_P |  |  |
| **D1** | *RNU6ATAC*<br>(NR_023344.1) | 9-134164529-A-C | n.36T>G | VUS | Variant in a<br>Gene of<br>Uncertain<br>Significance | Compound<br>Heterozygous | Autosomal<br>recessive |
|  |  | 9-134164537<br>-G-A | n.28C>T | VUS | Variant in a<br>Gene of<br>Uncertain<br>Significance |  |  |

**Table 2.** Phenotypic information for samples A1-D1. Information on the systems affected and clinical features seen in A1-D1. In the indicated individual, a + sign indicates the presence of a clinical feature, while a sign indicates the absence of a clinical feature.

| System Affected | Clinical Feature | A1 | B1 | C1 | C2 | D1 |
| --- | --- | --- | --- | --- | --- | --- |
| **Growth** | Short stature | + | + | + | + | + |
|  | Microcephaly | + | + | + | + | + |
| **Neurological** | Generalized hypotonia | + | + | - | - | + |
|  | Neurodevelopmental delay | - | + | - | - | + |
|  | Intellectual disability | + | + | - | - | + |
|  | Seizures | - | - | - | - | + |
|  | Ataxia | - | + | - | - | + |
|  | Ventriculomegaly | + | - | - | - | + |
|  | Peripheral neuropathy | - | + | + | - | - |
| **Musculoskeletal** | Scoliosis | + | - | - | - | - |
|  | Coxa valga | - | + | - | - | - |
|  | Syndactyly | + | - | - | - | + |
|  | Joint hypermobility | - | + | - | - | - |
| **Immunological** | Immunodeficiency | - | + | + | - | - |
| **Endocrine and Metabolic** | Growth hormone deficiency | + | - | + | - | - |
|  | Hypothyroidism | - | + | + | + | - |
|  | Adrenal insufficiency | - | - | + | + | - |
|  | Diabetes mellitus | - | - | + | - | - |
| **Ophthalmological** | Retinal Anomalies | + | + | - | + | - |
|  | Nystagmus | - | - | - | - | + |
|  | Oculomotor apraxia | - | - | - | - | + |
|  | Exotropia | + | - | - | - | - |
| **Cardiovascular** | Arrhythmia | + | - | - | - | - |
|  | Congestive heart failure | - | - | - | + | - |

Upon further investigation, all five of these cases were found to have rare, compound heterozygous variants in minor spliceosome snRNAs. Four of these cases (samples A1, B1, C1-2) were found to harbor rare, biallelic variants in *RNU4ATAC* **(Table 1)**. The mean number of MIGs with significant intron retention outliers in the cases with rare, biallelic variants in *RNU4ATAC* (175.8 (IQR [169 - 184.3])) was over 75 times higher than the mean number of MIGs with significant intron retention outliers in the rest of the whole blood cohort (2.3 (IQR [0 - 2])) **(Figure 5B)**. All four cases (A1-C2) shared MIGs with significant intron retention outliers with one another **(Figure S3, Table S2)**.

*RNU4ATAC* is transcribed into U4atac, which consists of six subregions, five of which are essential for the excision of minor introns. Crucial elements include stem I and stem II, which are important for base pairing with U6atac, *RNU6ATAC*’s transcribed counterpart. These regions are separated by a 5’ stem loop, which acts as a binding platform for proteins required for tri-snRNP formation and is also of high importance to minor splicing. Finally, the 3’ stem loop precedes a binding platform for Sm proteins, which are necessary for snRNA assembly and transport inside the nucleus. Some nucleotides in the 3’ stem loop are of variable importance to splicing and there is a fully dispensable region from position 92 to 105^70–74^.

One *RNU4ATAC* variant in A1 (NR_023343.3:n.13C>T) is in the stem II, and thus impacts a nucleotide of high importance to splicing, while the other is in the Sm protein binding region (NR_023343.3:n.116A>G) at a nucleotide that has been labeled of variable importance to minor intron excision. The variants seen in B1 are in the stem II (NR_023343.3:n.13C>T) and Sm protein-binding region (NR_023343.3:n.121T>G) and both nucleotides are of high importance to minor spliceosome function. Finally, one of the variants seen in both C1 and C2, who are siblings, is in the Sm protein-binding region (NR_023343.3:n.118T>C), and thus at a location of high importance to minor intron splicing. The other is in the 5’ stem loop (NR_023343.3:n.47T>G), but outside of the critical region and thus impacts a nucleotide of variable importance to minor intron splicing^70–74^ **(Figure 5C)**.

Finally, the phenotypes of the four aforementioned cases fit the expected clinical profile of individuals with an RNU4atac-opathy, defined here as someone with biallelic, rare, pathogenic, or likely pathogenic variants in *RNU4ATAC*. Biallelic pathogenic variants in *RNU4ATAC* are associated with a spectrum of autosomal recessive conditions referred to collectively as RNU4atac-opathy and were historically described as three conditions: microcephalic osteodysplastic primordial dwarfism type 1 (MOPD1; OMIM:210710)^75^, Roifman syndrome (RS; OMIM:616651)^76^, and Lowry-Wood Syndrome (LWS; OMIM:226960)^77^. Clinical features observed in cases A1-C2 include growth restriction, microcephaly, skeletal dysplasia, and intellectual disability^75–77^ **(Table 2, Table S3)**. Less common clinical features of RNU4atac-opathy were also noted, including immunodeficiency, as well as cardiac, ophthalmologic, and endocrine abnormalities. Our clinical and genetic findings recapitulate the known variability of phenotypes and genotypes associated with dysfunction of *RNU4ATAC*.

### Transcriptome-first approach identifies *RNU6ATAC* as a candidate disease gene

The remaining case (sample D1) with an excess of significant intron retention outliers in MIGs (z-score of 7.3, **Figure 5A**) had 153 MIGs impacted by significant intron retention outliers. This was over 80 times the number of significant intron retention outliers in MIGs observed in individuals without rare, biallelic variants in the minor spliceosome in the rest of the cohort (1.9 (IQR [0 - 2])) **(Figure 5B)**. Furthermore, D1 shared the vast majority of its genes with intron retention outliers (141 MIGs, 92.2%) with all four RNU4atac-opathy samples (Fisher’s exact test, p=2.72e-13) **(Figure 6A).** Only 12 MIGs with significant intron retention outliers were unique to this individual, within the range of unique outlier junctions for the RNU4atac-opathies (range 5-27, **Figure S3**).

**Figure 6.**
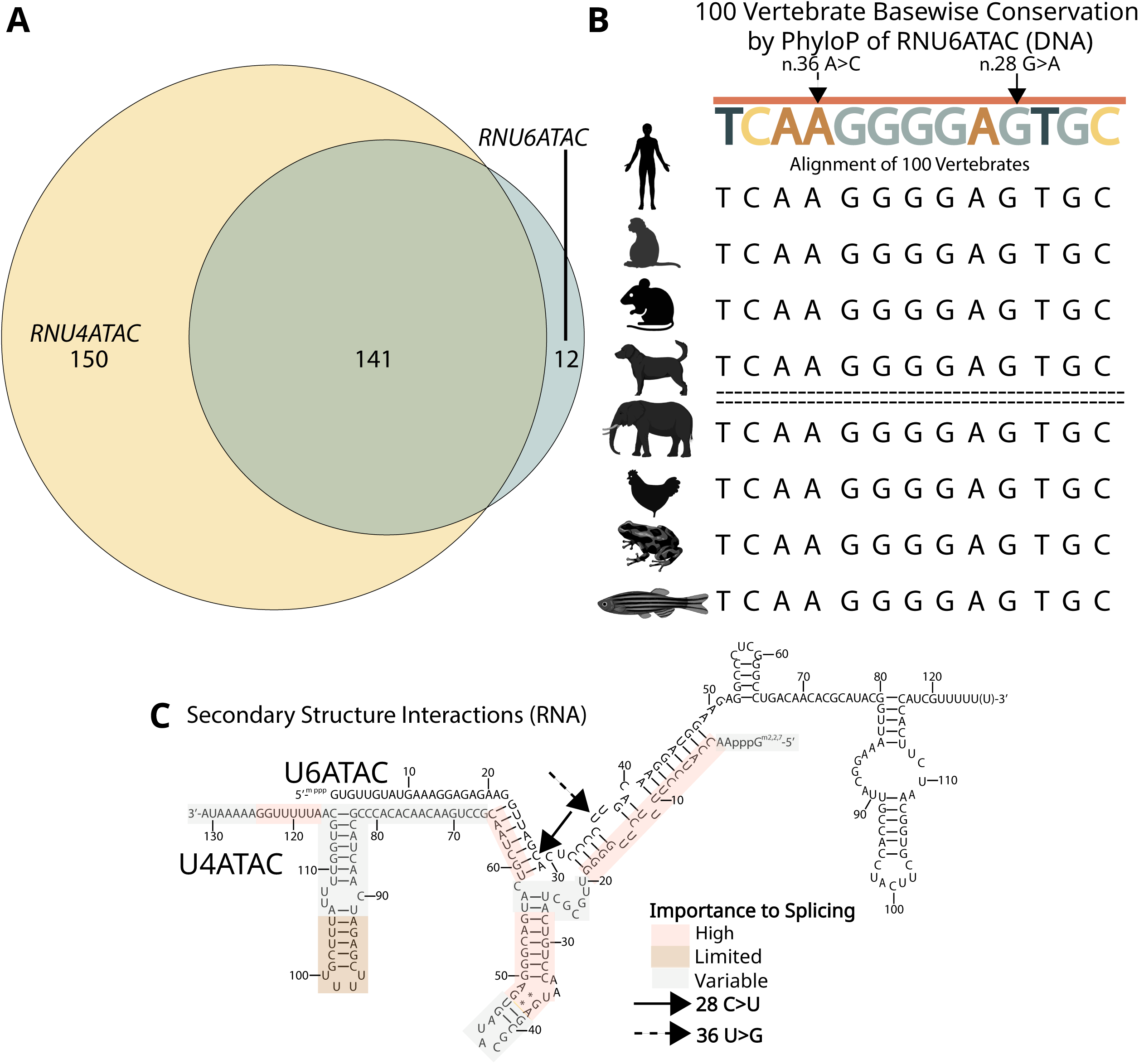
Outlier with excess intron retention events in minor intron containing genes (MIGs) found to harbor rare, conserved biallelic variants in *RNU6ATAC*. (A) Venn diagram of the genes shared between all four RNU4atac-opathy cases (A1-C2) and the case with rare, biallelic variants in *RNU6ATAC* (D1). All genes unique to the RNU4atac-opathy cases (A1-C2) are shown in yellow, while all genes unique to D1 (*RNU6ATAC* case) are shown in blue. All shared genes are shown in green. (B) Examination of conservation of RNU6ATAC (DNA). Conservation levels for each nucleotide were obtained using PhyloP to compare the nucleotides across 100 vertebrates. The brown line above the sequence indicates that all of the nucleotides in this region are highly conserved. The animals shown represent the following organisms: human, rhesus, mouse, dog, elephant, chicken, X. tropicalis, and zebrafish. The two RNU6ATAC variants seen in individual D1 are marked by arrows. (C) Secondary structure of the binding of U4ATAC and U6ATAC (RNAs) required for the formation of the catalytic splice site. The two variants in D1 are marked by the dashed and solid arrows. The relative importance of each nucleotide in RNU4ATAC to splicing is indicated in the colored legend.

No candidate variants in *RNU4ATAC* and *RNU12,* which are genes known to be involved with minor intron retention, were identified in D1. Given the extensive transcriptomic overlap between the RNU4atac-opathy cases (A1, B1, C1-2), and the remaining case with an excess of significant intron retention outliers in MIGs (D1), we expanded our analysis strategy and performed a targeted analysis of D1’s genome sequencing data to investigate rare variants (allele frequency <0.01, and no homozygous occurrence in gnomAD) in snRNAs of the minor spliceosome. This analysis identified rare, compound heterozygous variants in *RNU6ATAC* (NR_023344.1:n.36T>G and n.28C>T), a snRNA in the minor spliceosome^78,79^ **(Table 1)**.

Both *RNU6ATAC* variants identified in D1 are rare and present in the population at a frequency that is consistent with an autosomal recessive mode of inheritance. The n.36T>G variant is not reported in the gnomAD v4.1.0^80,81^ database and only one individual was found to have a heterozygous variant differing from the reference at that location (n.36T>A, allele frequency of 6.6e-6). The n.28C>T variant is present at a very low allele frequency of 7.9e-5 (12 out of 152,036 alleles) with no homozygous occurrences observed. Notably, the n.28C>T variant is the only variant differing from the reference at position 28 of the *RNU6ATAC* gene in gnomAD. Additionally, both variants are in a 39 base pair region in *RNU6ATAC* that is highly conserved^82,83^, and both are themselves highly conserved (PhyloP: 6.99 and 7.69^84^, **Figure 6B**).

The phenotype of case D1 includes intrauterine and postnatal growth restriction, microcephaly, epilepsy, intellectual disability, and ataxia. These clinical features resemble those observed in other conditions caused by pathogenic biallelic variants in minor spliceosome snRNAs^75–77^ and accordingly overlap with those seen in cases A1, B1, and C1-2 **(Table 2, Table S3)**.

The transcribed counterpart of *RNU6ATAC*, U6atac, extensively base-pairs with U4atac^70^, the transcribed counterpart of *RNU4ATAC,* to form the critical tri-snRNP along with U5^85^. It is thought that disruptions to the formation of this tri-snRNP contribute to the disease phenotypes seen in some individuals with RNU4atac-opathy^76^. The n.28C>T variant in D1 impacts a nucleotide that binds to U4atac’s stem I, which is known to be important to splicing^72,73,76,86^. The n.36C>T variant in D1 disrupts a tandem U formation in the *RNU6ATAC* transcript, which could destabilize the *RNU6ATAC* molecular structure^87^ or impact binding to nucleotides in *RNU4ATAC*’s stem II, which is another area of high importance to splicing^72,73,76,86^ **(Figure 6C).**

As U4atac and U6atac must successfully base-pair to form the critical tri-snRNP^72,73,76,86^, it is plausible that disruption of U6atac binding to U4atac could also cause Mendelian disease. These transcriptomic, genomic, and phenotypic findings suggest that our transcriptome-first approach followed by targeted genomic analysis of spliceosomal snRNAs identified a novel gene-disease candidate in *RNU6ATAC*. Importantly, the distinct transcriptomic signature may provide functional evidence supporting the pathogenicity of these variants. However, additional cases are needed to establish a formal gene-disease association. *RNU6ATAC* has been submitted through the GeneMatcher node of the Matchmaker Exchange, but no matches have been identified to date.

### Effect of sample size on results

Mertes et al., 2021 and Scheller et al., 2023 both indicate that the average number of outliers detected per sample increases along with sample size, but eventually plateau at sample sizes of 300 or more^41,42^. To examine the portability of our results to researchers and clinicians with access to fewer samples, we created six datasets consisting of 25, 50, 100, 150, 200, 300, and 400 samples, each of which contained no more than one sample with rare, biallelic variants in minor spliceosome snRNAs (A1).

Upon running each subset, we found that the number of significant intron retention outliers in MIGs in sample A1 increased significantly as the sample size increased (linear regression, p =0.047). Over two times the amount of significant intron retention outliers in MIGs were found in sample A1 using a sample size of 400 compared to 25 samples (168 vs 345). Additionally, the z-score associated with the number of significant intron retention outliers in MIGs in sample A1 increased significantly from 4.80 to 19.40 at sample sizes of 25 to 400, respectively (linear regression, p=7.3e-05, **Figure S4**).

### Use of Jaccard index for identifying minor spliceopathies

We sought to investigate whether FRASER2^42^ could identify the individuals with rare, biallelic variants in the minor spliceosome, known as minor spliceopathies. We found that all five cases with minor spliceopathies (A1-D1) had an excess of Jaccard index outliers in MIGs. Specifically, A1-D1 had z-scores of 4.9, 3.6, 3.5, 2.1, and 3.2 for the number of Jaccard index outliers in MIGs, respectively. Notably, using a z-score cut-off of 2 for the number of significant splicing outliers also identified two additional individuals, with z-scores of 2.1 and 2.1, as having an excess of Jaccard index outliers in MIGs **(Figure S5)**. For these individuals, we did not identify rare, biallelic variants in any of the snRNAs in the minor spliceosome.

We next analyzed the number of MIGs with Jaccard index outliers and found that cases A1-D1 had Z-scores of 5.2, 3.8, 3.6, 2.2, and 3.5, corresponding to 196, 148, 139, 91, and 137 MIGs impacted, respectively. Using this metric, no additional individuals with an excess of MIGs with Jaccard index outliers were identified **(Figure 5B)**. FRASER2^42^ analysis is thus also able to identify samples with the minor spliceosome transcriptional signature, but with less specificity.

## DISCUSSION

Transcriptomics is emerging as a complementary approach for rare disease diagnostics^9–16^. Current approaches using transcriptomics for rare disease diagnostics use a gene-centric approach, where the focus is on identifying variants resulting in *cis* outlier events, such as gene expression level or aberrant splicing^9–16^. The foundation of this approach has been numerous population studies that show enrichments for rare variants near transcriptome outlier events^17–22^. The majority of these studies exclude individuals with many transcriptome outliers and do not consider genes not previously attributed to human disease, as such outliers are assumed to be due to technical noise. While we found that splicing outliers were more likely to be in constrained disease-associated genes, these assumptions fail to acknowledge that global outlier events might instead be true biological signals of rare variants impacting key regulatory processes. Previous studies have implicated specific patterns of splicing dysregulation in the presence of pathogenic variants that disrupt splicing^31–33,37–40^. For example, distinct and cell-type specific patterns of aberrant splicing have been associated with repeat expansions in *DMPK*^88^, while pathogenic variants in *RBFOX1* are associated with aberrant splicing of exons ≤ 51 nucleotides in size^39,40^. In our cohort, we were able to identify individuals with excess splicing outliers and hypothesized that patterns of excess transcriptome-wide splicing outliers could aid in the discovery of causal variants in known spliceopathy-associated genes, as well as new gene-disease relationships.

One such pattern described by Olthof et al., 2021, showed that biallelic, pathogenic variants in minor spliceosome snRNAs *RNU4ATAC* and *RNU12* are associated with elevated intron retention in and around minor introns^32^. Given this information, we examined our cohort for individuals with excess intron retention in minor intron containing genes (MIGs). Using this approach, we identified five cases (samples A1, B1, C1-2, D1) with excess significant intron retention outliers impacting MIGs, all of which were identified as having biallelic variants in snRNAs critical to the minor spliceosome. Four of these individuals (samples A1, B1, C1-2) were found to have rare, biallelic variants in *RNU4ATAC.* All four samples had symptoms consistent with RNU4atac-opathy and did not harbor any additional rare variants of clinical interest, including in other minor spliceosome snRNAs. RNU4atac-opathy has a low diagnostic rate as the phenotype varies significantly depending on the specific variants involved^75–77^ and *RNU4ATAC* is located in the intron of the *CLASP1* gene, and therefore not routinely analyzed on clinical exome sequencing^89^. Based on our phenotypic, genetic, and transcriptomic findings, we are suggesting upgrading the classification of 4 *RNU4ATAC* variants and that this method could be used to provide supportive evidence for interpretation of variants of uncertain significance in *RNU4ATAC*.

We identified one additional case (sample D1) with an excess of significant intron retention outliers in MIGs with many phenotype features seen in individuals with pathogenic variants in the minor spliceosome, such as short stature, microcephaly, hypotonia, and intellectual disability^75–77^. Upon examination of their genome sequencing data, we found they harbored rare, compound heterozygous variants in *RNU6ATAC,* an snRNA in the minor spliceosome that hybridizes with *RNU4ATAC*^66,78^. Both variants are highly conserved^84^, at genomic positions with limited variation in the human population^80,81^, and could potentially disrupt the binding of *RNU6ATAC* to regions of high importance to splicing in *RNU4ATAC*^72,73,76,86^. The binding of *RNU4ATAC* to *RNU6ATAC* is crucial for the formation of the tri-snRNP^90^ and disruption of this binding has been associated with RNU4atac-opathy^91^. We hypothesize that the disruption of the binding of *RNU4ATAC* to *RNU6ATAC* in both RNU4atac-opathy cases and D1 could explain their shared transcriptomic and clinical features. However, the impact of the variants in D1 on the function of the minor spliceosome needs to be studied further to address this question. Overall, these genetic, transcriptomic, and phenotypic findings strongly suggest *RNU6ATAC* as a novel gene-disease candidate. While our transcriptomic-first approach can identify individuals with pathogenic variants in the minor spliceosome, we need additional samples and to perform functional validation to establish patterns specific to pathogenic variants within each snRNA.

In concordance with results from Scheller et al., 2023, we found that as sample size increased, the number of significant intron retention outliers detected in an RNU4atac-opathy case (A1) also increased. However, we show that batch-effects confound the number of splicing outlier junctions per individual, so combining cohorts might not always be a practical solution. We suggest best practices going forward should include the development of a batch-correction approach prior to outlier calling for splicing. Our blood cohort came from a heterogeneous processing background across three sequencing centers and four protocols. It is notable that despite these significant differences, the biological signature seen in cases with rare biallelic variants in *RNU4ATAC* or *RNU6ATAC* – excess significant intron retention outliers in MIGs – is robust to batch effects. We therefore expect this signature to be even stronger in samples that are uniformly processed.

Here we present an unbiased, transcriptomic-first approach for rare disease diagnosis and gene-disease relationship discovery in spliceopathies. While excess outlier patterns are influenced by technical or other covariates, disease-relevant excess outlier patterns are evident in rare disease cohorts. We highlight that investigating these biologically coherent patterns, such as intron retention outliers in MIGs, can increase the diagnostic yield for individuals with rare diseases. We further show that this approach can both inform variant curation and uncover new gene-disease relationships. As there are characteristic transcriptome-wide patterns of splicing dysregulation^31–33,37–40^ for some of the over 300 protein and RNA components involved in splicing^23,24^, we expect there may be other meaningful transcriptomic signatures. This is supported by recent discoveries of the roles of major spliceosome snRNAs, RNU4-2^92^ and RNU2-2p^93^, in neurodevelopmental diseases and reinforces the possibility that additional disease-causing variants that disrupt splicing remain to be uncovered. Combined, dissection of transcriptome-wide patterns of aberrant splicing represents a novel diagnostic approach for gene discovery and variant-to-functional interpretation of spliceopathies in rare disease patients.

## Data Availability

Our analyses and pipeline are fully available at https://github.com/maurermaggie/Transcriptome_Wide_Splicing_Analysis/tree/main. All RNA-seq data for samples enrolled in GREGoR are available in AnVIL through dbGap (phs003047.v1.p1). Most of the RNA-seq data for samples enrolled in the UDN is currently available through dbGap (phs001232.v6.p2). The remaining data will be uploaded to dbGaP as a part of the next UDN data freeze. Data is also available before the next freeze by a request to the author with evidence of dbGaP approval for UDN data.

## CONSORTIA

Members of the Undiagnosed Diseases Network: Alyssa A. Tran, Arjun Tarakad, Ashok Balasubramanyam, Brendan H. Lee, Carlos A. Bacino, Daryl A. Scott, Elaine Seto, Gary D. Clark, Hongzheng Dai, Hsiao-Tuan Chao, Ivan Chinn, James P. Orengo, Jennifer E. Posey, Jill A. Rosenfeld, Kim Worley, Lindsay C. Burrage, Lisa T. Emrick, Lorraine Potocki, Monika Weisz Hubshman, Richard A. Lewis, Ronit Marom, Seema R. Lalani, Shamika Ketkar, Tiphanie P. Vogel, William J. Craigen, Lauren Blieden, Jared Sninsky, Hugo J. Bellen, Michael F. Wangler, Oguz Kanca, Shinya Yamamoto, Christine M. Eng, Patricia A. Ward, Pengfei Liu, Adeline Vanderver, Cara Skraban, Edward Behrens, Gonench Kilich, Kathleen Sullivan, Kelly Hassey, Ramakrishnan Rajagopalan, Rebecca Ganetzky, Vishnu Cuddapah, Anna Raper, Daniel J. Rader, Giorgio Sirugo, Vaidehi Jobanputra, Allyn McConkie-Rosell, Kelly Schoch, Mohamad Mikati, Nicole M. Walley, Rebecca C. Spillmann, Vandana Shashi, Alan H. Beggs, Calum A. MacRae, David A. Sweetser, Deepak A. Rao, Edwin K. Silverman, Elizabeth L. Fieg, Frances High, Gerard T. Berry, Ingrid A. Holm, J. Carl Pallais, Joan M. Stoler, Joseph Loscalzo, Lance H. Rodan, Laurel A. Cobban, Lauren C. Briere, Matthew Coggins, Melissa Walker, Richard L. Maas, Susan Korrick, Jessica Douglas, Audrey Stephannie C. Maghiro, Cecilia Esteves, Emily Glanton, Isaac S. Kohane, Kimberly LeBlanc, Rachel Mahoney, Shamil R. Sunyaev, Shilpa N. Kobren, Brett H. Graham, Erin Conboy, Francesco Vetrini, Kayla M. Treat, Khurram Liaqat, Lili Mantcheva, Stephanie M. Ware, Breanna Mitchell, Brendan C. Lanpher, Devin Oglesbee, Eric Klee, Filippo Pinto e Vairo, Ian R. Lanza, Kahlen Darr, Lindsay Mulvihill, Lisa Schimmenti, Queenie Tan, Surendra Dasari, Adriana Rebelo, Carson A. Smith, Deborah Barbouth, Guney Bademci, Joanna M. Gonzalez, Kumarie Latchman, LéShon Peart, Mustafa Tekin, Nicholas Borja, Stephan Zuchner, Stephanie Bivona, Willa Thorson, Herman Taylor, Andrea Gropman, Barbara N. Pusey Swerdzewski, Camilo Toro, Colleen E. Wahl, Donna Novacic, Ellen F. Macnamara, John J. Mulvihill, Maria T. Acosta, Precilla D’Souza, Valerie V. Maduro, Ben Afzali, Ben Solomon, Cynthia J. Tifft, David R. Adams, Elizabeth A. Burke, Francis Rossignol, Heidi Wood, Jiayu Fu, Joie Davis, Leoyklang Petcharet, Lynne A. Wolfe, Margaret Delgado, Marie Morimoto, Marla Sabaii, MayChristine V. Malicdan, Neil Hanchard, Orpa Jean-Marie, Wendy Introne, William A. Gahl, Yan Huang, Aimee Allworth, Andrew Stergachis, Danny Miller, Elizabeth Blue, Elizabeth Rosenthal, Elsa Balton, Emily Shelkowitz, Eric Allenspach, Fuki M. Hisama, Gail P. Jarvik, Ghayda Mirzaa, Ian Glass, Kathleen A. Leppig, Katrina Dipple, Mark Wener, Martha Horike-Pyne, Michael Bamshad, Peter Byers, Sam Sheppeard, Sirisak Chanprasert, Virginia Sybert, Wendy Raskind, Nitsuh K. Dargie, Beth A. Martin, Chloe M. Reuter, Devon Bonner, Elijah Kravets, Holly K. Tabor, Jacinda B. Sampson, Jason Hom, Jennefer N. Kohler, Jonathan A. Bernstein, Kevin S. Smith, Matthew T. Wheeler, Meghan C. Halley, Page C. Goddard, Paul G. Fisher, Rachel A. Ungar, Raquel L. Alvarez, Shruti Marwaha, Terra R. Coakley, Euan A. Ashley, Ali Al-Beshri, Anna Hurst, Bruce Korf, Kaitlin Callaway, Martin Rodriguez, Tammi Skelton, Andrew B. Crouse, Jordan Whitlock, Mariko Nakano-Okuno, Matthew Might, William E. Byrd, Changrui Xiao, Eric Vilain, Jose Abdenur, Kathyrn Singh, Rebekah Barrick, Sanaz Attaripour, Suzanne Sandmeyer, Tahseen Mozaffar, Albert R. La Spada, Elizabeth C. Chao, Maija-Rikka Steenari, Alden Huang, Brent L. Fogel, Esteban C. Dell’Angelica, George Carvalho, Julian A. Martínez-Agosto, Manish J. Butte, Martin G. Martin, Naghmeh Dorrani, Neil H. Parker, Rosario I. Corona, Stanley F. Nelson, Yigit Karasozen, Aaron Quinlan, Alistair Ward, Ashley Andrews, Corrine K. Welt, Dave Viskochil, Erin E. Baldwin, John Carey, Justin Alvey, Laura Pace, Lorenzo Botto, Nicola Longo, Paolo Moretti, Rebecca Overbury, Russell Butterfield, Steven Boyden, Thomas J. Nicholas, Matt Velinder, Gabor Marth, Pinar Bayrak-Toydemir, Rong Mao, Monte Westerfield, Brian Corner, John A. Phillips III, Kimberly Ezell, Lynette Rives, Rizwan Hamid, Serena Neumann, Ashley McMinn, Joy D. Cogan, Thomas Cassini, Alex Paul, Dana Kiley, Daniel Wegner, Erin McRoy, Jennifer Wambach, Kathy Sisco, Patricia Dickson, F. Sessions Cole, Dustin Baldridge, Jimann Shin, Lilianna Solnica-Krezel, Stephen Pak, Timothy Schedl, Hector Rodrigo Mendez, Brianna Tucker, Beatriz Anguiano, Mia Levanto, Suha Bachir, Laurens Wiel, Stephen B Montgomery, Tanner D Jensen, John E. Gorzynski, Sara Emami, Laura Keehan, Jennifer Schymick, Taylor Maurer, Alexander Miller, Andres Vargas, Amanda M. Shrewsbury, Bianca E. Russell, Layal F. Abi Farraj, Elizabeth A Worthey, Tarun KK Mamidi, Brandon M Wilk, Rachel Li, Jennifer Morgan, Chun-Hung Chan, Paul Berger, Mohamad Saifeddine, Isum Ward, Jason Schend, Megan Bell, Dr. Francisco Bustos velasq, Taylor Beagle, Miranda Leitheiser, Runjun Kumar, Donald Basel, Michael Muriello, Brett Bordini, Michael Zimmermann, Abdul Elkadri, James Verbsky, Julie McCarrier

Members of the Genomics Research to Elucidate the Genetics of Rare Diseases (GREGoR) consortium: Siwaar Abouhala, Kaileigh Ahlquist, Miguel Almalvez, Emily Alsentzer, Raquel Alvarez, Mutaz Amin, Kailyn Anderson, Peter E. Anderson, Euan Ashley, Themistocles Assimes, Light Auriga, Christina Austin-Tse, Michael J. Bamshad, Rebekah Barrick, Rebekah Barrick, Samantha Baxter, Sairam Behera, Shaghayegh Beheshti, Gill Bejerano, Sami Belhadj, Seth Berger, Jon Bernstein, Sabrina Best, Benjamin Blankenmeister, Elizabeth E. Blue, Krista Bluske, Eric Boerwinkle, Emily Bonkowski, Devon Bonner, Philip M. Boone, Philip M. Boone, Leandros Boukas, Denver Bradley, Harrison Brand, Kati J. Buckingham, Daniel Calame, Colleen Carlston, Jennefer Carter, Silvia Casadei, Lisa Chadwick, Clarisa Chavez, Ziwei Chen, Yong-Han Cheng, Ivan Chinn, Jessica X. Chong, Zeynep Coban-Akdemir, Andrea J. Cohen, Sarah Conner, Matthew P. Conomos, Karen Coveler, Ya Allen Cui, Zain Dardas, Colleen P. Davis, Moez Dawood, Ivan de Dios, Celine de Esch, Celine de Esch, Emmanuèle Délot, Salil Deshpande, Stephanie DiTroia, Harsha Doddapaneni, Haowei Du, Michael Duyzend, Michael Duyzend, Michael Duyzend, Iman Egab, Evan E. Eichler, Sara Emami, Ivy Evergreen, Jamie Fraser, Vincent Fusaro, Mira Gandhi, Vijay Ganesh, Brandon Garcia, Kiran Garimella, Richard Gibbs, Sophia B. Gibson, Casey Gifford, Carmen Glaze, Pagé Goddard, Stephanie Gogarten, Nikhita Gogate, William W. Gordon, John E. Gorzynski, William Greenleaf, Christopher Grochowski, Emily Groopman, Emily Groopman, Rodrigo Guarischi-Sousa, Sanna Gudmundsson, Jonas (A. Gus) Gustafson, Stacey Hall, Caitlin Harrington, John Harting, William T. Harvey, Sohaib Hassan, Megan Hawley, Benjamin D. Heavner, Martha Horike-Pyne, Yun-Hua Hsiao, Jianhong Hu, Yongqing Huang, Karan Jaisingh, Minal Jamsandekar, Gail P. Jarvik, Tanner Jensen, Shalini Jhangiani, David Jimenez-Morales, Christopher Jin, Aimee Juan, Ahmed K. Saad, Jessica Kain, Rachid Karam, Laura Keehan, Rupesh Kesharwani, Charles Hadley King, Julia Klugherz, Arthur Ko, Anshul Kundaje, Soumya Kundu, Samuel M. Lancaster, Katie Larsson, Arthur Lee, Gabrielle Lemire, Jesse Levine, Richard Lewis, Wei Li, Yidan Li, Pengfei Liu, Bojan Losic, Jonathan LoTempio, James (Jim) Lupski, Jialan Ma, Daniel MacArthur, Annelise Y. Mah-Som, Medhat Mahmoud, Brian Mangilog, Dana Marafi, Sofia Marmolejos, Daniel Marten, Eva Martinez, Colby T. Marvin, Shruti Marwaha, F. Kumara Mastrorosa, Dena Matalon, Taylor Maurer, Susanne May, Sean R. McGee, Lauren Meador, Heather C. Mefford, Hector Rodrigo Mendez, Olfa Messaoud, Alexander Miller, Danny E. Miller, Stephen Montgomery, Yulia Mostovoy, Mariana Moyses, Mariana Moyses, Chloe Munderloh, Donna Muzny, Ashana Neale, Sarah C. Nelson, Matthew B. Neu, Thuy-mi P. Nguyen, Jonathan Nguyen, Robert Nussbaum, Emily O’Heir, Melanie O’Leary, Briana O’Leary, Sebastian Ochoa Gonzalez, Jeren Olsen, Ikeoluwa Osei-Owusu, Anne O’ÄôDonnell-Luria, Miranda P.G. Zalusky, Evin Padhi, Lynn Pais, Piyush Panchal, SHRUTI PANDE, Karynne E. Patterson, Sheryl Payne, Davut Pehlivan, Davut Pehlivan, Paul Petrowski, Alicia Pham, Georgia Pitsava, Astaria/Sara Podesta, Elizabeth Porter, Jennifer Posey, Jaime Prosser, Guanghao Qi, Wanqiong Qiao, Thomas Quertermous, Archana Rai, Heidi Rehm, Chloe Reuter, Matthew A. Richardson, Andres Rivera-Munoz, Lindsay Romo, Oriane Rubio, Kathryn Russell, Aniko Sabo, Monica Salani, Monica Salani, Kaitlin Samocha, Alba Sanchis-Juan, Sarah Savage, Stuart Scott, Evette Scott, Adriana E. Sedeño-Cortés, Fritz Sedlazeck, Jillian Serrano, Gulalai Shah, Ali Shojaie, Moriel Singer-Berk, Mugdha Singh, Riya Sinha, Joshua D Smith, Kevin Smith, Hana Snow, Michael Snyder, Kayla Socarras, Olivia M. Sommerland, Lea M. Starita, Brigitte Stark, Sarah Stenton, Sarah Stenton, Andrew B. Stergachis, Adrienne Stilp, Suchitra Sudarshan, V. Reid Sutton, V. Reid Sutton, Jui-Cheng Tai, Jui-Cheng Tai, Michael Talkowski, Christina Tise, Catherine C. Tong, Philip Tsao, Rachel Ungar, Grace VanNoy, Eric Vilain, Gaby Villard, Isabella Voutos, Kim Walker, Juliana Walrod, Chia-Lin Wei, Ben Weisburd, Jeffrey M Weiss, Chris Wellington, Ziming Weng, Lauren Westerfield, Emily Westheimer, Marsha Wheeler, Matthew Wheeler, Laurens Wiel, Michael Wilson, Monica Wojcik, Issac Wong, Frank Wong, Quenna Wong, Changrui Xiao, Jiaoyang Xu, Rachita Yadav, Rachita Yadav, Yao Yang, Qian Yi, Jiye Yu, Bo Yuan, Christina Zakarian, Jianhua Zhao, Jimmy Zhen

## ACKNOWLEDGEMENTS

We extend thanks to all research participants in the UDN and GREGoR consortia. We would also like to thank members of the Montgomery lab, the GREGoR consortium, and the Stanford Genetics Department who gave invaluable feedback and assistance throughout this entire project. From the Stanford site, we would like to specifically thank Chloe Reuter, Jennefer Carter, Laurens van de Wiel, Daniel Nachun, and Kate Lawrence. We would like to acknowledge the individuals at the Utah UDN site, especially Ashley Andrews, Lorenzo Botto, and Erin Baldwin, who acquired several samples included in this project.

This work required computing resources from the Stanford Genetics Bioinformatics Service Center (supported by NIH Instrumentation Grant S10 OD025082) and would not have been possible without the Stanford SCG cluster administrators, specifically Chris Jeon and Karl Kornel. Several figures were made with BioRender.

Research reported in this manuscript was partly funded by the National Human Genome Research Institute at the National Institutes of Health, as part of GREGoR consortium, through Grant Nos. U01HG011762 and U01HG011755. This publication was also supported by the Undiagnosed Diseases Network, which was aided by the NIH Common Fund through the Office of the NIH Director, the National Institute of Neurological Disorders and Stroke, and the Office of Strategic Coordination as part of Grant Nos. U01HG010217 and U01HG010218. M.T.A. was further supported by the National Science Foundation Graduate Research Fellowship under Grant No. DGE-2146755. V. S. G. is supported by the NIH NIAMS K23AR083505, and the BroadIgnite Award. The content of this manuscript does not necessarily represent the official views of the National Institutes of Health or the National Science Foundation and is solely the responsibility of the authors.

## AUTHORSHIP CONTRIBUTION

V. S. G., A. O., M. T. W., S. B. M., and J. A. B. oversaw the design and implementation of the study. M. T. A., R. A. U, P. C. G., and A. M. M. performed the initial transcriptomic quality control and alignment steps. M. T. A. also set up and ran the outlier detection tools FRASER and FRASER2 on all samples, as well as evaluated the results and performed all statistical analyses, which can be found on the github repository she set up and maintained. She created all figures, table S1, and table S2, as well as wrote all sections of the paper except the “Variant Classification” and “Genome Sequencing” sections, as well as sentences related to A1-D1’s phenotype, which R. M. and D. E. B wrote. R. M. oversaw the inclusion of A1, C1, and C2, diagnosed B1 with RNU4atac-opathy, and identified the *RNU6ATAC* variants in D1. He provided phenotypic descriptions for A1-D1, created Tables 1, 2, and S2 (which were formatted by M. T. A.), and assessed and classified the genetic variants in both *RNU4ATAC* and *RNU6ATAC*. He also coordinated inter-institutional collaboration/ sharing of RNA-seq data with UDN sequencing cores, which M. T. A., A. M. M., J. V. N., S. M., G. B., S. B., and M. T. aided in. R. A. U, E. M. P., A. M. M., and J. M. provided computational support. R. A. U also provided support in writing, organization, experimental design, and figure creation, the latter two of which E. M. P. also aided in. D. E. B., D. M., G. L., and R. M. performed recruitment of cohort members and provided access and interpretation of patient data. K. S. S., S. A. S., L. L., Z. N., and G. B. performed wet lab analyses, and K. S. S., S. A. S, and G. B. all provided information vital to the “Methods” section. G. B., S. B., and M. T. provided patient samples and phenotype information from the Miami UDN site. All authors edited and reviewed the manuscript.

