## Supplemental Methods for "Transcriptome-wide outlier approach identifies individuals with minor spliceopathies"

#### Genome Sequencing

Genome sequencing for cases A1, C1, C2, and D1 were performed by Baylor Genetics, through the Undiagnosed Disease Network (UDN), using methods previously described by Splinter et al., 2018[1]. In brief, libraries were prepared using a PCR-free 550-bp insert size protocol by the Hyper Prep kit. Sequencing was performed using Illumina NovaSeq 6000 platform for 150 bp paired-end reads. The SNPTrace Panel from the Fluidigm SNPtype platform was performed as a quality control measure.  Data analysis and interpretation were performed using the Illumina Dragen pipeline. FastQ data were aligned to the human reference genome build GRCh38 using the Illumina Dragen BioIT Platform while variant calling on the BAM file was performed using the Illumina Dragen haplotype-based variant calling system.

Genome sequencing for case B1 was performed by the Stanford Clinical Genomics Service Lab, through the GREGoR Stanford Site. Genome sequencing library preparation was performed using the KAPA Hyper Prep Kit (Roche Sequencing and Life Science, Indianapolis, IN) and KAPA Unique-Dual Indexed (UDI) adapters (Roche Sequencing and Life Science) according to manufacturer instructions. In brief, ~500 ng of genomic DNA was fragmented to ~400-450 bp and subjected to end repair and A-tailing, followed by adapter ligation, bead-based purification, and size selection. Adaptor-ligated libraries were quantified by qPCR and pooled for sequencing, which was performed on the NovaSeq 6000 (Illumina, San Diego, CA) using either S2 or S4 flow cells and 150 bp paired-end sequencing according to manufacturer instructions. Genome sequencing depth of coverage was targeted to ≥40X. Sequencing data was analyzed using the Stanford Clinical Genomics genome pipeline. Demultiplexed FASTQ files were generated from binary base call (BCL) files using bcl2fastq (https://emea.support.illumina.com/sequencing/sequencing_software/bcl2fastq-conversion-software.html), and sequencing reads were mapped to GRCh38. Variant calling was performed using the DRAGEN Germline Pipeline (Illumina), which employs a Germline Small Variant Caller for SNVs/indels, and the Structural Variant (SV) caller (Manta) and CNV Caller for structural variants.

### TABLES

#### Table S1. Gene sets used for enrichment analyses

| Gene set Description | URL Source | Source |
|---|---|---|
| **Haploinsufficient** | https://github.com/maurermaggie/Transcriptome_Wide_Splicing_Analysis/blob/main/Gene_Information/haploinsufficient.tsv | ClinGen dataset (Cormier et al., 2021[2]) |
| **Autosomal recessive** | https://github.com/maurermaggie/Transcriptome_Wide_Splicing_Analysis/blob/main/Gene_Information/autosomal_recessive.tsv | Blekhman et al., 2008[3]; Berg et al., 2013[4] |
| **Autosomal dominant** | https://github.com/maurermaggie/Transcriptome_Wide_Splicing_Analysis/blob/main/Gene_Information/autosomal_dominant.tsv | Blekhman et al., 2008[3]; Berg et al., 2013[4] |
| **Olfactory receptor** | https://github.com/maurermaggie/Transcriptome_Wide_Splicing_Analysis/blob/main/Gene_Information/olfactory_receptors.tsv | Mainland, et al., 2015[5] |
| **CRISPR non-essential** | https://github.com/maurermaggie/Transcriptome_Wide_Splicing_Analysis/blob/main/Gene_Information/CRISPR_nonessential_genes.tsv | Hart et al., 2017[6] |
| **Developmental delay** | https://www.ebi.ac.uk/gene2phenotype/downloads/DDG2P.csv.gz | Firth et al., 2011[7]; Fitzgerald, et al., 2015[8]; Wright et al., 2015[9]; McRae, et al., 2017[10]; Wright, et al., 2018[11] |
| **OMIM** | https://omim.org/downloads/ | OMIM dataset (Amberger et al., 2019[12]) |

**Table S1. Gene sets used for enrichment analyses.** Information on the gene sets used for the enrichment analysis of the outliers detected using FRASER and FRASER2. The haploinsufficient, autosomal recessive, autosomal dominant, olfactory receptor, and CRISPR non-essential gene sets can also be found at the Macarthur GitHub at: https://github.com/macarthur-lab/gene_lists/tree/master/lists.

### Table S2. Step-wise comparison of the MIGs with intron retention events shared between A1-D1

| Sample: Sample Comparison | Adjusted P-value | Odds Ratio |
|---|---|---|
| **A1:B1** | 1.51e-20 | 7.38 |
| **A1:C1** | 1.19e-11 | 4.11 |
| **A1:C2** | 1.05e-12 | 4.60 |
| **B1:C1** | 1.25e-6 | 2.72 |
| **B1:C2** | 1.63e-6 | 2.75 |
| **C1:C2** | 1.47e-36 | 21.4 |
| **D1:A1-C2** | 2.72e-13 | 7.72 |

**Table S2. Step-wise comparison of the MIGs with intron retention events shared between A1-D1.**
Comparison of the MIGs with intron retention events shared, in a step-wise fashion, between all possible combinations of A1-D1. The adjusted p-values and odds ratio were calculated by Fisher's exact test[13] **(Methods)**.

Table S3. Human Phenotype Ontology terms for samples A1-D1

| Family-Case ID | Human Phenotype Ontology (HPO) Terms |
|---|---|
| A1 | Short stature [HP:0004322], Growth hormone deficiency [HP:0034323], Microcephaly [HP:0000252], Generalized hypotonia [HP:0001290], Intellectual disability [HP:0001249], Scoliosis [HP:0002650], Ventriculomegaly [HP:0002119], Finger syndactyly [HP:0006101], Pigmentary retinopathy [HP:0000580], Exotropia [HP:0000577], Fused labia minora [HP:0000063], Arrhythmia [HP:0011675] |
| B1 | Short stature [HP:0004322], Microcephaly [HP:0000252], Neurodevelopmental delay [HP:0012758], Hypotonia [HP:0001252], Intellectual disability, mild [HP:0001256], Ataxia [HP:0001251], Peripheral neuropathy [HP:0009830], Immunodeficiency [HP:0002721], Hypothyroidism [HP:0000821], Peripheral retinal degeneration [HP:0007769], Coxa valga [HP:0002673], Joint hypermobility [HP:0001382] |
| C1 | Short stature [HP:0004322], Microcephaly [HP:0000252], Gait disturbance [HP:0001288], Peripheral neuropathy [HP:0009830], Diabetes mellitus [HP:0000819], Hypothyroidism [HP:0000821], Adrenal insufficiency [HP:0000846], Growth hormone deficiency [HP:0034323], Immunodeficiency [HP:0002721] |
| C2 | Short Stature [HP:0004322], Microcephaly [HP:0000252], Rod-cone dystrophy [HP:0000510], Hypothyroidism [HP:0000821], Adrenal insufficiency [HP:0000846], Congestive heart failure [HP:0001635] |
| D1 | Short Stature [HP:0004322], Microcephaly [HP:0000252], Generalized hypotonia [HP:0001290], Global developmental delay [HP:0001263], Intellectual disability [HP:0001249], Seizures [HP:0001250], Ataxia [HP:0001251], Nystagmus [HP:0000639], Oculomotor apraxia [HP:0000657], Syndactyly [HP:0001159], Ventriculomegaly [HP:0002119] |

**Table S3. Phenotypic information for samples A1-D1.**
Information on the phenotypes seen in samples A1-D1, given as Human Phenotype Ontology (HPO) terms.
