## Supplementary material for "Transcriptome-wide outlier approach identifies individuals with minor spliceopathies": Figure S1

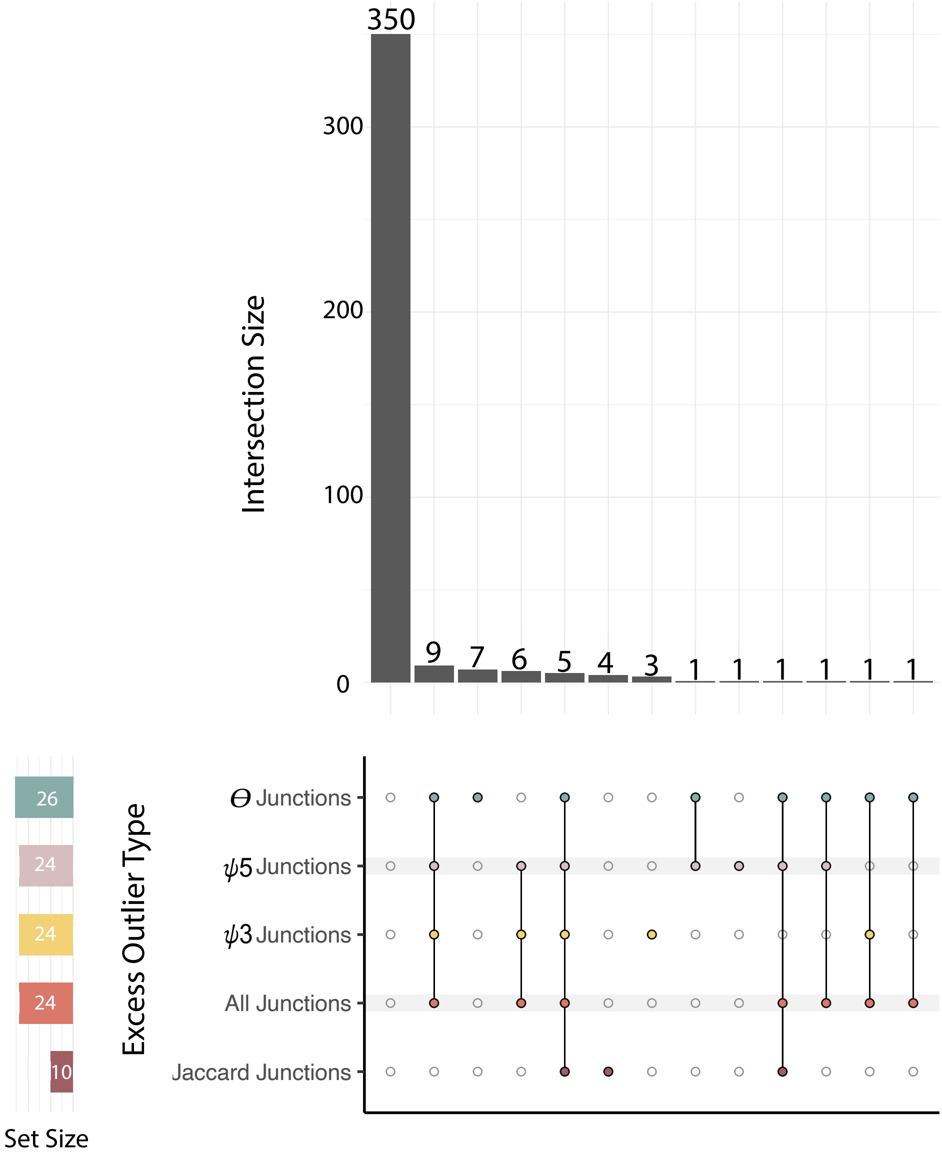


**Supplemental Figure 1. Sharing of samples with excess outliers across different FRASER and FRASER2 metrics**

Upset plot showing the number of samples with an excess of significant outliers across one metric that also have an excess of significant outliers across different metric(s) of FRASER or FRASER2. No significance patterns (* for $q \leq 0.05$, ** for $q \leq 0.01$, *** for $q \leq 0.001$, and *** for $q \leq 0.0001$) are shown as all correlations are not significant using logistic regression and false discovery rate correction. "All Junctions" refers to samples having an excess number of FRASER outliers when the number of significant ψ5, ψ3, and θ outliers are summed together.
