## Supplementary material for "Transcriptome-wide outlier approach identifies individuals with minor spliceopathies": Figure S2

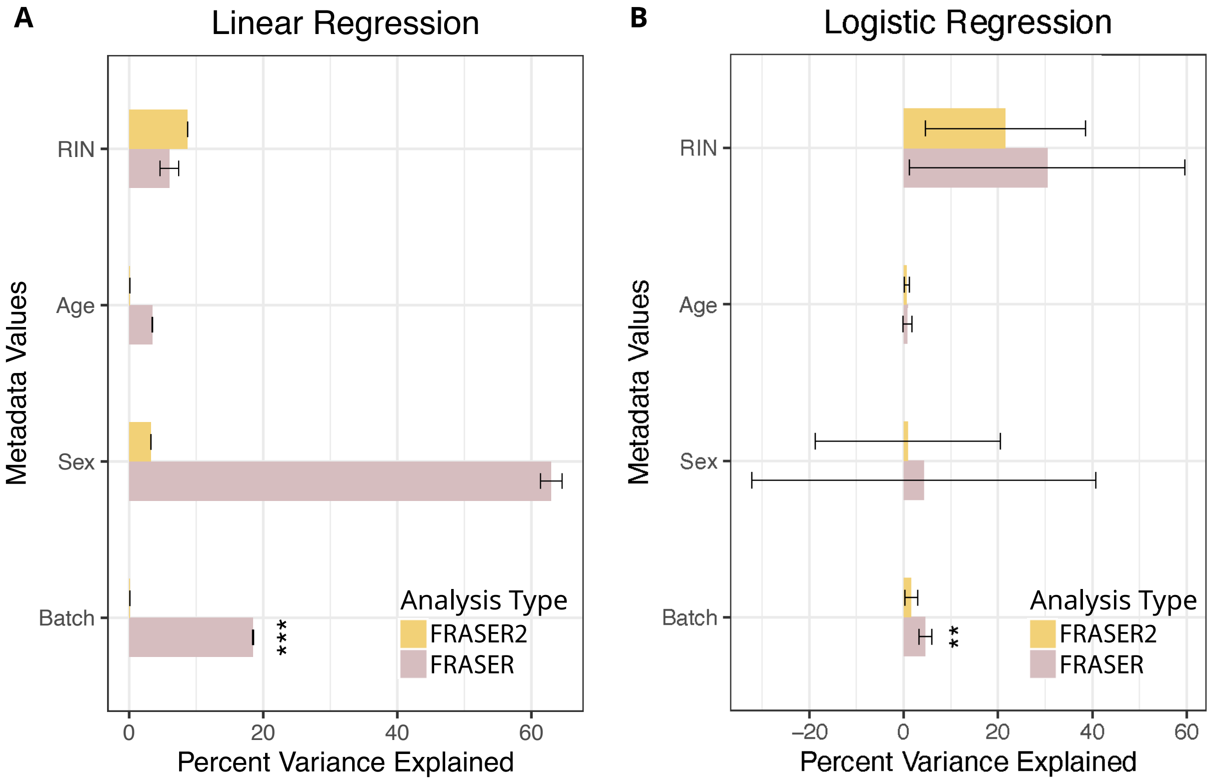


**Supplemental Figure 2. Association between technical and biological covariates and number of significant outlier junctions, outlier status**

Bar plots showing the percent variance explained by four technical and biological covariates. Percent variance was taken from the estimate calculated by a linear regression model (A) or logistic regression model (B), with error bars corresponding to the standard error. Significance levels were calculated after false discovery rate correction (* for q ≤ 0.05, ** for q≤ 0.01, *** for q ≤ 0.001, and *** for q ≤ 0.0001).

(A) Bar plots showing the percent variance explaining the total number of significant (q < 0.05 and $|\Delta\psi|$ <= 0.3) outlier junctions detected by FRASER2 (top, yellow) and FRASER (bottom, purple) per person.

(B) Bar plots showing the percent variance explaining the likelihood of an individual having an excess of splicing outliers. Logistic regression of the covariates was applied to a field containing either a 1 or a 0 for each individual. For the top yellow bars, 1 indicated the individual had an excess number of FRASER2's Jaccard index outlier junctions, while for the bottom purple bars, 1 indicated the individual had an excess of significant outlier junctions as calculated by any metric of FRASER ($\psi 5$, $\psi 3$, $\theta$, and combined). For both analyses (top and bottom), 0 indicated they did not have an excess of significant outlier junctions for their respective metrics.
