## Supplementary material for "Transcriptome-wide outlier approach identifies individuals with minor spliceopathies": Figure S3

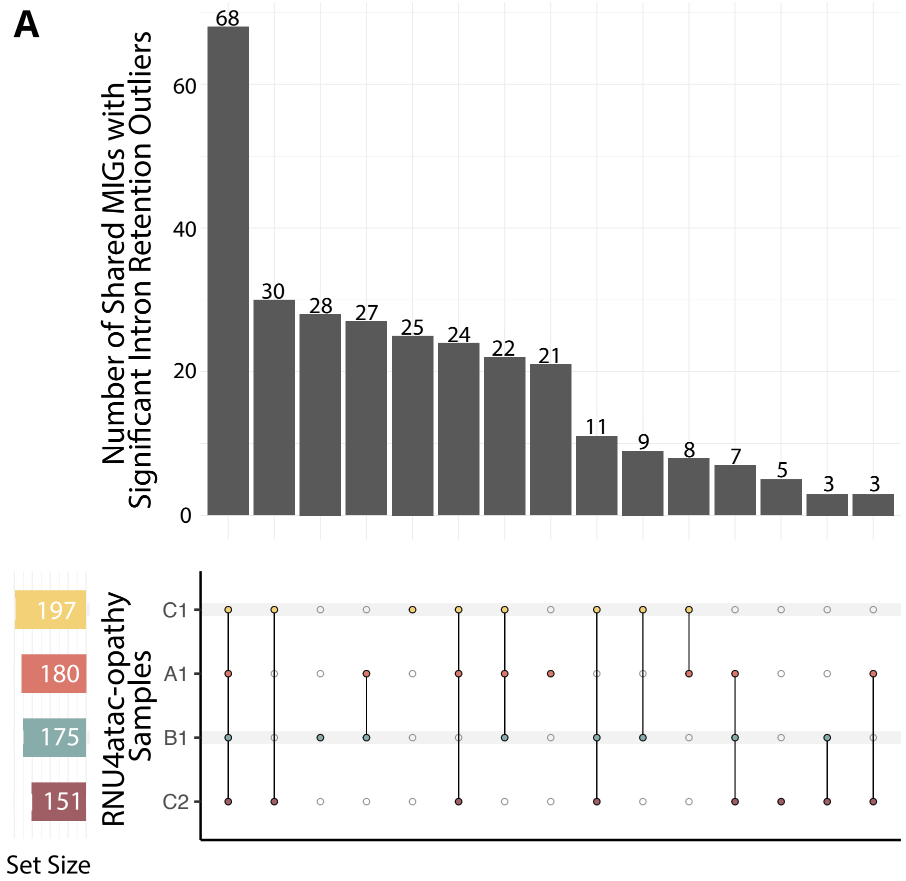


**Supplemental Figure 3. Minor intron containing genes (MIGs) with significant intron retention outliers shared between RNU4atac-opathy cases A1-C2**

(A) Upset plot showing the number of MIGs with significant intron retention outliers shared between the RNU4atac-opathy cases (A1- C2). All pairwise relationships between cases A1-C2 are significant (**Supplemental Table 2**).
