## Supplementary material for "Transcriptome-wide outlier approach identifies individuals with minor spliceopathies": Figure S4

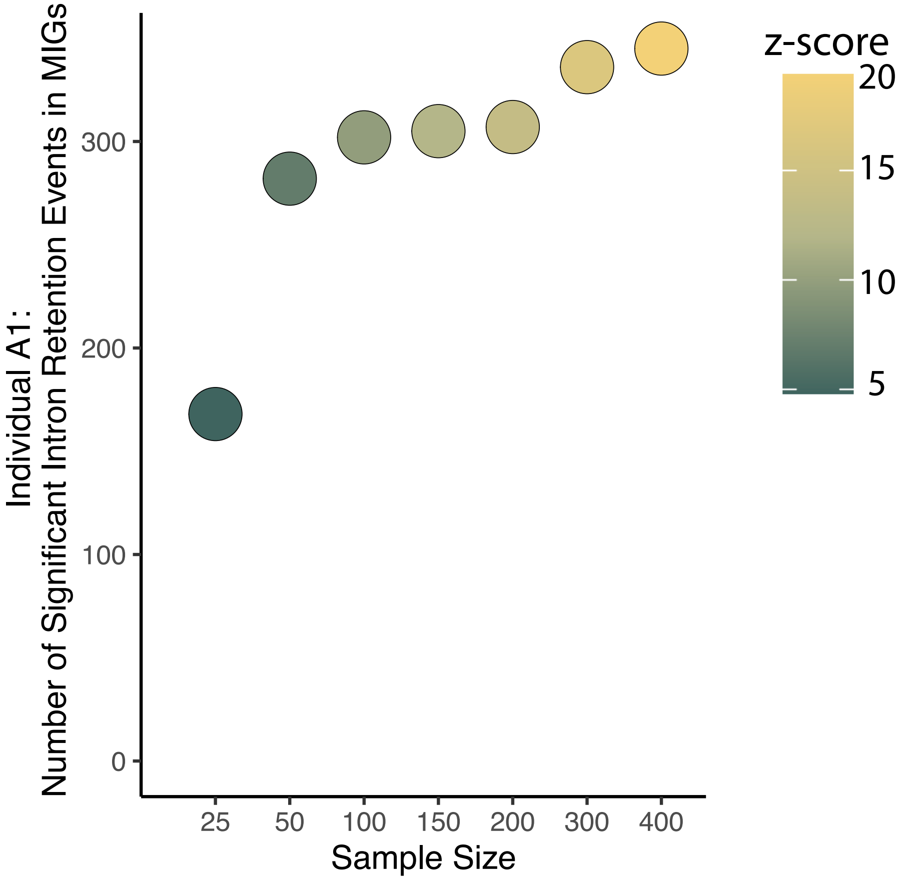


**Supplemental Figure 4. Sample size impacts the number of significant intron retention events detected in minor intron containing genes (MIGs) for RNU4atac-opathy case A1**

Plot depicting how changes in sample size, shown on the X-axis, impact the number of significant intron retention events in MIGs detected by FRASER in individual A1, shown on the Y-axis. The z-score of individual A1 relative to the rest of the subsampled cohort is indicated by the color of the circle, which ranges from dark green at 25 on the X-axis (corresponding to a z-score of 4.8) to bright yellow at 400 on the X-axis (corresponding to a z-score of 19.4). All other samples with rare, biallelic variants in the minor spliceosome were removed from the cohort and then the subsampled cohorts were chosen iteratively. Cohort 25 and the subsequent additions to each cohort (for example, 25 random samples were added to cohort 25 to make cohort 50) were chosen randomly after the removal of samples B1-D1. The z-score was calculated by subtracting the subsampled cohort mean from the number of significant intron retention events in MIGs in A1 and then dividing the resulting number by the standard deviation of the subsampled cohort.
