## Supplementary material for "Transcriptome-wide outlier approach identifies individuals with minor spliceopathies": Figure S5

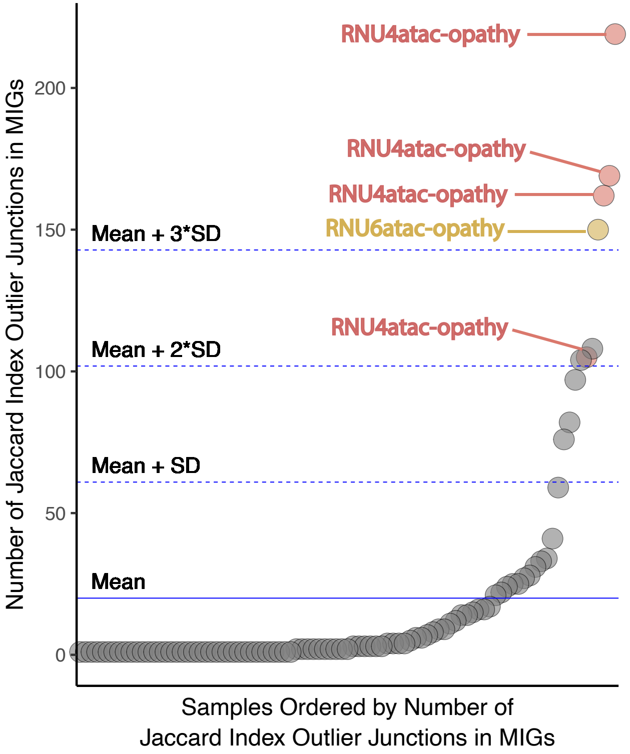


**Supplemental Figure 5. Outliers with excess significant Jaccard index outliers in minor intron containing genes (MIGs) identified in individuals with rare, biallelic variants in minor spliceosome snRNAs *RNU4ATAC* and *RNU6ATAC***

Plot showing the number of significant Jaccard index outliers in MIGs. Each dot represents an individual, with the position on the Y-axis representing the number of significant Jaccard index outliers in MIGs detected in that individual. The X-axis is ordered by the number of significant Jaccard index outliers in MIGs detected per individual. Individuals with rare, biallelic variants in *RNU4ATAC* are the salmon-colored dots labeled as “RNU4atac-opathy”, while the individual with rare, biallelic variants in *RNU6ATAC* is represented by the mustard-colored dot labeled as “RNU6atac-opathy.” Gray dots do not have rare, biallelic variants in any snRNA components of the minor spliceosome.
